# Multimodal Image Guidance in Subthalamic Deep Brain Stimulation for Parkinson’s Disease

**DOI:** 10.1101/2025.09.03.25334924

**Authors:** Patricia Zvarova, Christina van der Linden, Ningfei Li, Konstantin Butenko, Thea Berger, Garance M. Meyer, Ilkem Aysu Sahin, Lukas L. Goede, Bahne H. Bahners, Barbara Hollunder, Till A. Dembek, Andrew R. Pines, Martin Reich, Jens Volkmann, Vincent J.J. Odekerken, Rob M. A. de Bie, Xin Xu, Zhipei Ling, Chen Yao, Andrea A. Kühn, Surjo R. Soekadar, Kerstin Ritter, Michael T. Barbe, Veerle Visser-Vandewalle, Michael D. Fox, Jan Niklas Petry-Schmelzer, Nanditha Rajamani, Andreas Horn

**Affiliations:** Movement Disorder and Neuromodulation Unit, Department of Neurology, Charité-Universitätsmedizin Berlin, corporate member of Freie Universität Berlin and Humboldt-Universität zu Berlin, Berlin, Germany; Einstein Center for Neurosciences Berlin, Charité – Universitätsmedizin Berlin, Berlin, Germany; Center for Brain Circuit Therapeutics, Department of Neurology, Brigham and Women’s Hospital, Harvard Medical School, Boston MA 02115, USA; Network Stimulation Institute, Department of Stereotactic and Functional Neurosurgery, University Hospital Cologne, Germany; Department of Neurology, Faculty of Medicine and University Hospital Cologne, University of Cologne, Cologne, Germany; Department of Neurology, Center for Movement Disorders and Neuromodulation, Medical Faculty and University Hospital Düsseldorf, Heinrich Heine University Düsseldorf; Institute of Clinical Neuroscience and Medical Psychology, Medical Faculty and University Hospital Düsseldorf, Heinrich Heine University Düsseldorf; Berlin School of Mind and Brain, Humboldt-Universität zu Berlin, Berlin, Germany; Brigham & Women’s Hospital, Harvard Medical School, Department of Psychiatry; Department of Neurology, University Clinic of Würzburg, Josef-Schneider-Str. 11, 97080, Würzburg, Germany; Department of Neurology, Amsterdam University Medical Center, University of Amsterdam, Amsterdam, The Netherlands; Department of Neurosurgery, Chinese PLA General Hospital, Beijing, 100853, China; Department of Neurosurgery, Hainan Hospital of Chinese PLA General Hospital, Sanya, Hainan, 572000, China; Department of Neurosurgery, The National Key Clinic Specialty, Shenzhen Key Laboratory of Neurosurgery, the First Affiliated Hospital of Shenzhen University, Shenzhen Second People’s Hospital, Shenzhen, 518035, China; Bernstein Center for Computational Neuroscience Berlin, Berlin, Germany; NeuroCure Clinical Research Centre, Charité – Universitätsmedizin Berlin, corporate member of Freie Universität Berlin and Humboldt-Universität zu Berlin, Berlin, Germany; Clinical Neurotechnology Laboratory, Department of Psychiatry and Neurosciences (CCM), Charité - Universitätsmedizin Berlin, Berlin, Germany; Berlin Center for Advanced Neuroimaging (BCAN), Charité— Universitätsmedizin Berlin, Berlin, Germany; Hertie Institute for AI in Brain Health, University of Tübingen, Germany; Department of Stereotactic and Functional Neurosurgery, Faculty of Medicine and University Hospital Cologne, University of Cologne, Cologne, Germany; MGH Neurosurgery & Center for Neurotechnology and Neurorecovery (CNTR) at MGH Neurology Massachusetts General Hospital, Harvard Medical School, Boston, MA 02114, USA

**Author notes:** Correspondence to: Patricia Zvarova, Movement Disorders and Neuromodulation Section Charité – Universitätsmedizin Berlin, Charitéplatz 1, 10117 Berlin, Germany. These authors contributed equally to this work (Nanditha Rajamani, Andreas Horn).

**Keywords:** Parkinson’s disease, Subthalamic nucleus, Deep brain stimulation, Connectomics, Neuroimaging, Outcome predictions

## Abstract

**Background:** Accurate electrode placement and individual stimulation parameters influence the outcomes of subthalamic deep brain stimulation in Parkinson’s disease. Neuroimaging-based models can help evaluate how electrode placement impacts improvement, aiming to reduce the burden of DBS programming. However, most existing models have been developed to explain differences between patients rather than differences between contacts within the same patient, leaving the clinical relevance of image-guided programming unclear.

**Methods:** We analyzed data from patients with Parkinson’s disease treated with subthalamic deep brain stimulation to develop and validate a neuroimaging-informed model of motor improvement measured by the Unified Parkinson’s Disease Rating Scale. Five approaches were tested: active contact coordinates, electric fields, tract activations, as well as structural and functional networks. All approaches were integrated into a combined ridge regression model and validated using two hold-out datasets.

**Results:** The sample included 236 patients (604 stimulation sites), divided into a training cohort (N = 129), a retrospective validation cohort (N = 89), and a prospectively acquired validation cohort (N = 21 electrodes). Consistent with expectations, our model explained approximately 11% of the variance in unseen group-level data (*R^2^* = 0.11, *p* = 0.001). At the individual level, the model identified the optimal clinical contact or its neighboring contact in all but one case (mixed-effects *R^2^*= 0.32, *p* = 1 x 10^-16^).

**Conclusion:** The imaging-informed model explained the expected variance at the group level and demonstrated potential for guiding stimulation programming, suggesting that image-guided approaches may improve clinical decision-making while reducing the need for lengthy postoperative testing.

## Introduction

Deep brain stimulation (DBS) is an effective treatment option for Parkinson’s disease (PD), but achieving the best outcomes requires precise electrode placement and carefully selected stimulation parameters.^1^ DBS programming identifies the electrode contact or combination of contacts that provide the greatest therapeutic effect. In most centers, this process is time-consuming and burdensome, often requiring multiple follow-up visits, especially during the first year after surgery.^2^ Several strategies have been proposed to streamline this process, including electrophysiological sensing^3,4^ and image-guided programming.^5,6^ Image guidance maps the electrode’s location relative to brain anatomy,^7^ and may be informed by probabilistic “sweet-spots”,^8^ optimal sets of structural connections,^9,10^ or polysynaptic whole-brain network targets.^11^ First randomized controlled clinical trials have shown that image-guided programming can achieve non-inferior results compared to expert-based DBS programming.^6^

The performance of image-guided models has often been evaluated by their ability to explain variance in clinical outcomes across unseen group-level cohorts.^9,11–13^ While this provides insight into DBS mechanisms and helps explain some variance in clinical outcomes, applying such models in a clinical setting requires accurate predictions on the individual level, posing fundamentally different challenges. In clinical practice, these models can be applied to suggest suitable electrode contacts or stimulation parameters to optimize care in individual patients. Consequently, some studies have already quantified how well algorithm- recommended contacts align with those selected by expert clinicians. Importantly, the ultimate aim in this case is to match expert-based decisions or even to improve outcomes in individual patients.^5,6,9^ Although they are related, these two tasks are distinct goals. Explaining group-level variance and improving individual patient outcomes each presents different challenges to image-guidance models.

The first task (explaining group-level outcomes) is complicated by the many factors that influence DBS outcomes beyond active contact placement (assuming the electrode reaches the target nucleus). In our focused literature review we identified numerous sources of heterogeneity in clinical outcomes after DBS, including disease subtypes, duration, structural and anatomical variations, comorbidities, and more. We must emphasize that these considerations are specific to subthalamic nucleus DBS (STN-DBS) for PD, and may largely differ for other indications.^14,15^ Here, group-level data often suffers from inter- and intra-rater variability in clinical scoring and suboptimal clinical imaging, especially when aggregated across multiple centers (Figure 1A). While some studies compared the impact of several factors, no single study exhaustively quantified their relative contributions in a combined analysis. This introduces an unclear covariance structure, which requires assumptions about factors such as disease subtype or imaging quality (e.g., due to head motion). These assumptions introduce uncertainty and potential bias. Although it remains unclear how much of the variance in clinical response image-guidance models can explain in heterogeneous (“noisy”) group-level data, based on our literature review, we hypothesize that approximately 10% of the variance in clinical outcomes in such data can be attributed to electrode placement.^9^ A more detailed discussion of this analysis may be found in the Supplementary material (Supplementary Text 1 and Supplementary Table 1).

**Figure 1.**
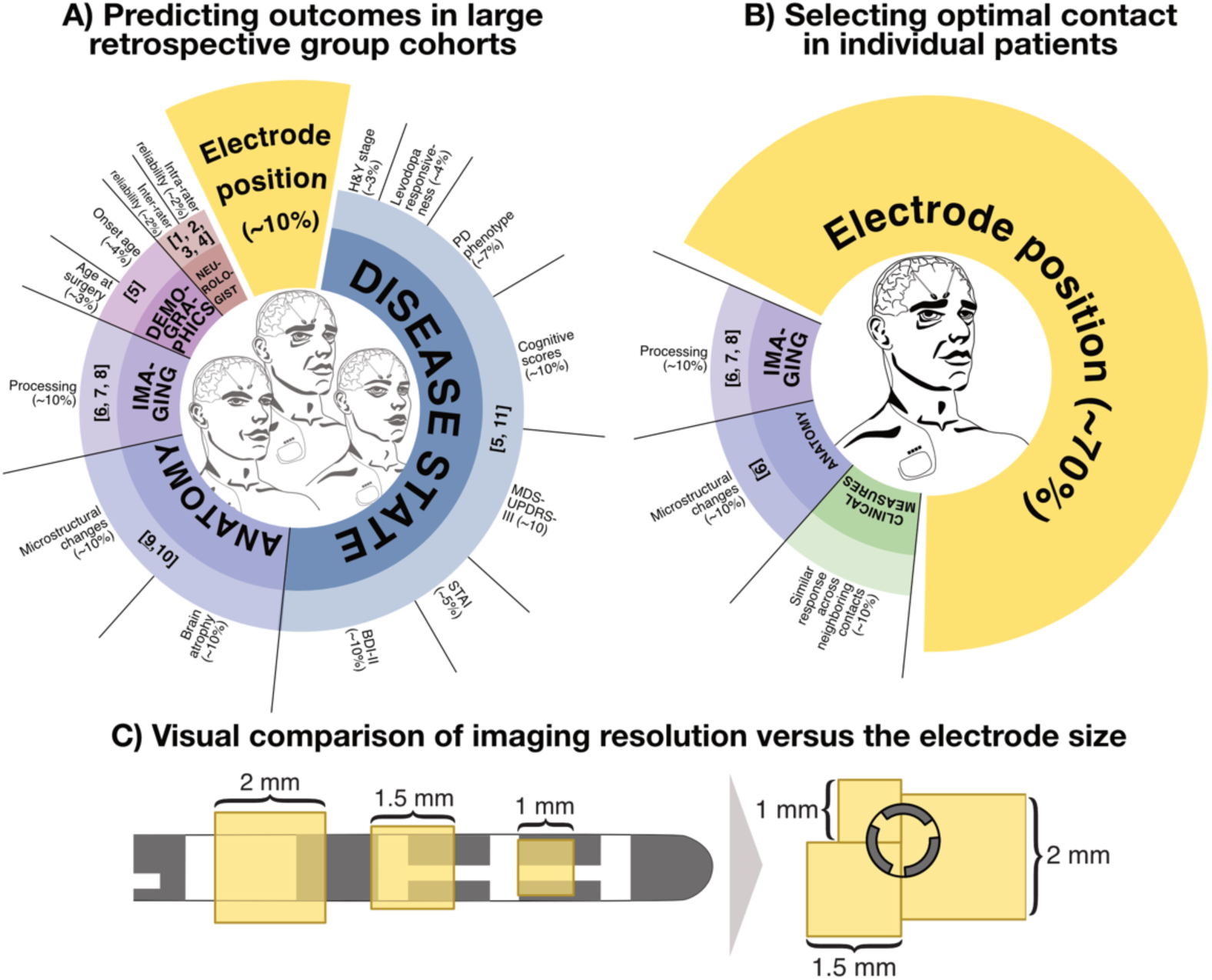
Factors influencing long-term clinical outcomes and accuracy of STN-DBS image guidance models. A) Our focused literature search suggested that we lack conclusive evidence on how various factors complicate the task of estimating clinical improvements across heterogeneous group-level STN-DBS datasets in the treatment of Parkinson’s disease (PD). The identified contributing factors include disease state, macro- and microscopical anatomical brain abnormalities, imaging*, demographics, and examiner/neurologist). Based on estimates from the literature, we expected the electrode position to explain approximately 10% of the variance in clinical improvements (considering it reaches the defined target nucleus). The numbers in the figure were approximated, based on the factors deemed influential in the literature (see Supplementary Text 1 and Supplementary Table 1 for source details). The innermost circle includes the specific category, the middle circle includes the reference number (see Supplementary Table 1), the outermost circle includes the specific factor. B) When DBS imaging guidance tools are tasked with suggesting optimal DBS contacts in individual patients, the structure of nuisance variables differs significantly. Numerous noisy/heterogeneous factors populating group-level data (A) are eliminated, because the model must select options within the same patient, same brain, and same subthalamic nucleus. However, we estimated that image guidance models could still be affected by microstructural anatomical changes. For instance, perivascular (Virchow-Robin) spaces located closer to some of the contacts would lead to changes in outcomes that the guidance model cannot predict. Furthermore, the task is nontrivial to begin with, since neighboring electrode contacts will often lead to comparable clinical responses, i.e., there may not always be an unequivocal “top contact”, leading to some degree of ambiguity in the data. We therefore attributed an additional 10% of the variance to such clinical measurement uncertainty. Lastly, variance arising from imaging data processing remains consistent across both group-level and individual-level modeling scenarios. Therefore, while optimal contact selection is not the sole contributor to clinical improvement, we hypothesize it explains a substantial 70% of the variance in clinical improvement in a single patient. All references can be found in the Supplementary material (Supplementary Text 1 and Supplementary Table 1). C) Whether explaining variance in clinical improvement at the group level, or suggesting optimal stimulation contacts in an individual patient, imaging resolution remains a key limiting factor, especially when making choices across individual contacts on a single electrode. The required resolution may theoretically exceed that of imaging currently used in clinical practice. For example, resolution in typical structural MRI is around 1 mm^3^, in diffusion MRI between 1-2 mm^3^ and in functional MRI between 2-3 mm^3^. The spacing between the centers of adjacent contacts on a typical DBS electrode is about 2 mm, with segmented contacts being placed even closer together. As a result, a single voxel may often cover multiple DBS contacts, highlighting the challenges imaging resolution poses for the accuracy of image-guided DBS programming. H&Y stage: Hoehn and Yahr Scale; MDS-UPDRS-III: MDS Unified Parkinson’s Disease Rating Scale; STAI: State Trait Anxiety Inventory; BDI-II: Beck Depression Inventory *It is important to note that no study has deliberately tested for imaging processing or reconstruction errors. However, multiple reports speak of reconstruction errors displacements of at least 1 mm. To our knowledge, the only study that tested for artificially introduced spatial uncertainty imposed a jitter of N = 258 electrode placements using a 3D Gaussian distribution with a 2 mm full width half maximum. Models created after spatial jittering correlated with the original model at approximately R = 0.8, corresponding to 36% non-shared variance between the original and jittered models. However, only a fraction of this unexplained variance can be attributed to neuroimaging-related spatial uncertainty, as additional factors such as random noise or imperfect model fitting are also expected to contribute. Therefore, we estimate that neuroimaging-related uncertainty accounts for approximately 9% of the variance in clinical improvement following DBS.^8^

The second task (selecting optimal stimulation parameters in an individual patient) eliminates many of the confounding variables that populate the first task. For instance, patient-specific characteristics such as PD subtype, disease duration, and age stay constant for any given patient. Additionally, the same clinician may assess responses to stimulation at each contact within a single session, eliminating observer bias (Figure 1B). However, this individualized approach introduces novel challenges that are different in nature. Namely, contacts along the same electrode are merely millimeters apart from one another, and neighboring contacts will often produce comparable stimulation results. Therefore, identifying a single best contact may be limited by imaging resolution, especially when segmented electrodes are used (Figure 1C). Furthermore, microstructural anatomical variations unknown to the model, such as Virchow-Robin spaces, may still cause unexpected clinical outcomes, even with optimal electrode placement and contact selection.^16^ Nonetheless, if image-guided models are truly used to inform DBS programming, these challenges must be overcome. Based on these considerations (see Supplementary Text 1 for details), we hypothesized that image guidance models would have clear clinical value in informing programming decisions for individual patients.

To empirically test these theoretical considerations, we developed a multimodal image- guided model based on a large multicenter cohort and evaluated its performance in the two aforementioned tasks: (1) estimating and explaining variance in clinical outcomes in an independent group-level cohort, and (2) suggesting optimal stimulation contacts for individual patients. After constructing the model, our first aim involved testing it on between- subject variance. Our second aim sought to improve clinical efficiency, as identifying a subset of optimal clinical contact(s) could significantly reduce DBS programming time.^6^ Within this effort, we constructed and validated the image-guided model informed by five distinct methods of analysis, namely anatomical coordinates, “sweet spots”, optimal tract connections and distributed structural and functional networks, using data from 236 patients (604 stimulation sites) across five international DBS centers.

## Materials and methods

### Patient cohorts, imaging and clinical assessment

We collected an overall sample of 247 patients with bilateral STN-DBS treated for PD from six different cohorts. Of these, data from 236 patients were retained for the analyses (Supplementary Figure 1). Informed consent was obtained in accordance with the Declaration of Helsinki, and the study methods have been approved by the Institutional Review Board at Charité-Universitätsmedizin Berlin. The discovery dataset consisted of 129 retrospectively enrolled patients from three centers. The first validation dataset included two additional retrospective cohorts comprising 89 patients. The second validation dataset consisted of 18 patients in whom the clinical response of each contact was tested at 2 mA (*N* = 21 electrodes) as part of clinical trial (DRKS00026596) at the University Hospital Cologne.

Clinical symptoms (off medication) before and after surgery were assessed using the total motor score of the Unified Parkinson’s Disease Rating Scale (UPDRS-III) for all patients, except the contact-wise stimulation cohort, in which upper limb contralateral hemi-scores were acquired based on the rigidity, finger tapping, resting tremor, postural tremor and kinetic tremor items of the MDS-UPDRS III, rated by a clinician who was blinded to the current stimulation settings. The contact-wise stimulation cohort was derived during the routine monopolar review process. A full MDS-UPDRS-III assessment was impractical due to time constraints. The unilateral focus of this cohort also reflects the decision to limit testing to the hemisphere contralateral to the tremor-affected upper extremity.

### DBS electrode localization and electric field calculation

DBS electrode location was reconstructed using Lead-DBS v3.1 software run in MATLAB R2022b (The MathWorks Inc., Natick, MA, USA), following an advanced, state-of-the-art protocol.^1^ Pre- and post-operative images were first linearly co-registered using Advanced Normalization Tools (ANTs)^17^ followed by non-linear normalization into the ICBM 2009b Nonlinear Asymmetric (“MNI”) template space using ANTs SyN,^18^ and SPM, as implemented in Lead-DBS.^1^ Normalization misalignments were corrected using WarpDrive.^19^ Potential intraoperative brain shift was corrected using subcortical refinements of the registration.^20^ In the case of post-operative CT (*N* = 201) images, the electrode placements were reconstructed using the phantom-validated Precise and Convenient Electrode Reconstruction for Deep Brain Stimulation (PaCER) approach.^21^ Electrodes from post-operative MRI scans (*N* = 46) were reconstructed using the TRAC/CORE algorithm. If necessary, results from both methods were manually inspected and refined.^22^ The DiODe algorithm^23^ was used to estimate the rotational orientation of directional leads, followed by manual inspection and adjustment when needed.

Stimulation volumes were approximated by calculating electric field magnitudes (E-fields) using OSS-DBS v2.^24^ E-field calculation approximates voltage gradients administrated from the electrode to voxels in space, yielding higher values closer to its source and lower values with increased distance from the source. OSS-DBS was used via its integration into Lead-DBS, and the modelling details can be found in Butenko et al.^24^ Stimulation volumes were estimated in native space and warped to template space using the same non-linear registration field described earlier.

### Statistical analysis

#### Models associated with the optimal clinical improvement

Models that associated stimulation sites with clinical improvements on four levels were calculated based on the discovery cohort (*N* = 129) using five different methods. First, at the electrode level, active contact locations were averaged per hemisphere and weighted by their improvement. This yielded a single “optimal” coordinate. Second, at the stimulation volume level, we correlated patient-specific E-fields with clinical improvements using established DBS Sweet Spot Mapping.^13^ For the third and fourth level, we applied DBS Fiber Filtering^25^ and DBS Network Mapping^11^ to identify optimal tract, structural network and functional network targets, following established methods. All details are provided in the Supplementary material (Supplementary Methods 1).

#### Validation of the models and calculation of surrogate values

Model performance at all levels was first assessed in a circular (in-sample) manner, then evaluated using 10-fold cross-validation (CV) within the discovery dataset. Models on each level (coordinates, E-fields, tracts and structural/functional networks) generated estimates that were optimized to be strongly correlated with the real clinical improvement for each patient. These estimated values did not directly predict UPDRS-III improvement but rather represented surrogate variables (or scores) that would optimally be correlated with UPDRS-III improvement values. For example, on the coordinate level, the surrogate for the clinical scores was expressed by proximity of the patient specific chronic contact to the optimal coordinate. The closer the stimulation site to the target, the better the outcome would be, according to the model. For the remaining methods, surrogate variables represented Pearson’s correlations between the model and patient profile (for more details see Supplementary Methods 2). Because these surrogate values were calculated from the entire discovery dataset, this was a circular (in-sample) analysis. Next, the procedure was repeated using a 10-fold CV design, applied analogously at the coordinate, E-field, tract, and network levels. Using the coordinate level as an example, the discovery dataset was split into 10 folds. The optimal stimulation coordinates were iteratively calculated based on nine folds, and proximity to the resulting coordinate was measured in the left-out tenth fold. This procedure was repeated for each left-out fold. This ensured the data from the estimated (hold-out) fold were not included in the optimal stimulation target calculation, preventing circularity. This cross-validation was carried out across all levels, strictly taking care that no data leakage was possible in any of the processing steps. Note that we deliberately refrain from providing p-values for in-sample analyses throughout the manuscript, but we provide p-values when models were subjected to cross-validation (circumventing circularity).

#### Combined ridge regression model of clinical improvement

All five models relied on DBS electrode localization, which can lead to high covariance across the model estimates. However, prior research suggests that combining different modeling strategies may still explain additional outcome variance.^11^ Each method likely has unique pitfalls that can lead to false predictions in individual cases. We hypothesized that combining all five strategies into a joint model could enhance robustness and better generalize to unseen data by reducing the risk of overfitting to method-specific noise. To do so, we fit ridge regression (using a regularization parameter of *λ* = 50, see Supplementary Methods 3 for more details) to the full training dataset using the in-sample surrogate values from all five models. Ridge regression was chosen to address multicollinearity, reduce overfitting, and increase predictive power on the out-of-sample validation dataset. Multicollinearity of our scores arose not only from overlapping anatomical or functional information across different modalities, but also from shared variance introduced by our preprocessing pipelines.

#### Validation of the combined model using an independent dataset

Each of the five models was tested on a previously unseen, heterogeneous (hold-out) test dataset from two different centers and a prospectively acquired contact-wise stimulation cohort. Model-derived surrogate variables for clinical improvement in the test cohort were generated using models trained exclusively on the discovery dataset. Specifically, Euclidean distance and Pearson’s correlation coefficients were calculated in the same way as in the discovery cohort. These model-derived surrogates were then used to (1) test the performance of each model individually across the five levels, and (2) test the combined performance using the regression model.

First, the ridge regression model, trained on the normalized surrogate variables of the discovery cohort, was used to predict clinical outcomes in the retrospective validation dataset.

Importantly, this was done without refitting the model to the validation dataset in any way; that is, the *ß* coefficients were calculated exclusively based on the discovery cohort.

The combined ridge regression model was also evaluated using prospectively acquired patients who were stimulated using a fixed amplitude of 2 mA on each of the eight contacts. Here, each electrode had eight clinical outcome values (one for each contact) and was treated as an individual dataset. As in the retrospective validation cohort, each contact received five model-derived surrogate values (proximity to the optimal coordinate and various Pearson’s correlation coefficients with the sweet spot, optimal tract, structural and functional network profiles). These five surrogates were then used to derive clinical improvement in the test cohort, based on the beta estimates from the ridge regression model trained exclusively on the discovery cohort.

To assess predictive performance,^26^ we calculated the coefficient of determination (R^2^) as R^2^ = 1 – Residual sum of squares / total sum of squares for the first out-of-sample validation cohort.

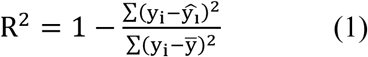

Where ∑(*y_i_* − *ŷ_i_*)^2^ is the residual sum of squares and ∑(𝑦_i_ − *ȳ*)^2^ is the total sum of squares between the predicted improvements and empirical improvements.

## Data availability

All code used to analyze the dataset is openly available within Lead-DBS (including sweet spot mapping, fiber filtering and network mapping software: https://github.com/leaddbs/leaddbs).

Cohort-wise demographic and clinical outcomes from the training and the test cohorts are available in Supplementary Table 2 and 3. We cannot openly share patient imaging data due to data sharing and privacy regulations, but they can be made available upon request to the corresponding primary investigator. The corresponding author and the principal investigator (PZ and AH) commit to returning data requests within a time frame of 30 days.

## Results

### Patient demographics and clinical results

The discovery dataset included 129 PD patients with STN-DBS from three cohorts (Berlin = 51, Würzburg = 44, Amsterdam = 34). The retrospective, heterogeneous group-level validation dataset included two cohorts (Beijing = 41, Würzburg = 48). The second validation cohort included 18 prospectively acquired contact-wise stimulation data (21 electrodes). Clinical scores, demographics and active contact placements are provided in the Supplementary material (Supplementary Table 2, Supplementary Table 3, and Supplementary Figure 4).

### Optimal DBS target sites at various levels of analyses

Optimal target engagement was measured at the coordinate, stimulation volume, tract, and structural and functional network levels in the discovery dataset. As expected, the optimal coordinate and the sweet spot were found in the dorsolateral motor STN. Optimal structural connectivity profiles peaked in the supplementary motor area (SMA), pre-supplementary motor area (pre-SMA), and superior frontal gyrus, while functional connectivity profiles were anticorrelated with the primary motor cortex (M1) (Supplementary Results 1 and Supplementary Figure 5). At the coordinate level, the proximity of each patient’s active contact significantly correlated with clinical improvements (in-sample: *R* = 0.39; 10-fold CV: *R* = 0.37, *p* < 0.001). This indicates that the closer the active contact to the optimal stimulation coordinate, the higher the improvement (Figure 2A). At the sweet-spot level, the spatial correlation between each patient’s E-field and the optimal sweet-spot profile also led to significant results (in-sample: *R* = 0.42; ten-fold CV: *R* = 0.39, *p* < 0.001; Figure 2B). At the tract level, patients whose stimulation overlapped with the optimal tract profiles had better clinical outcomes (in sample: *R* = 0.43; ten-fold CV: *R* = 0.36, *p* < 0.001; Figure 2C). At the network level, stimulation profiles that were more closely correlated with the identified structural (structural in-sample: *R* = 0.36; ten-fold CV: *R* = 0.41, *p* < 0.001) and functional networks (functional in-sample: *R* = 0.37; ten-fold CV: *R* = 0.36, *p* < 0.001) yielded higher clinical outcomes (Figure 2D and 2E). Overall, the similarity between in-sample and cross-validated R-values suggests a minimal degree of overfitting in any of these models.

**Figure 2.**
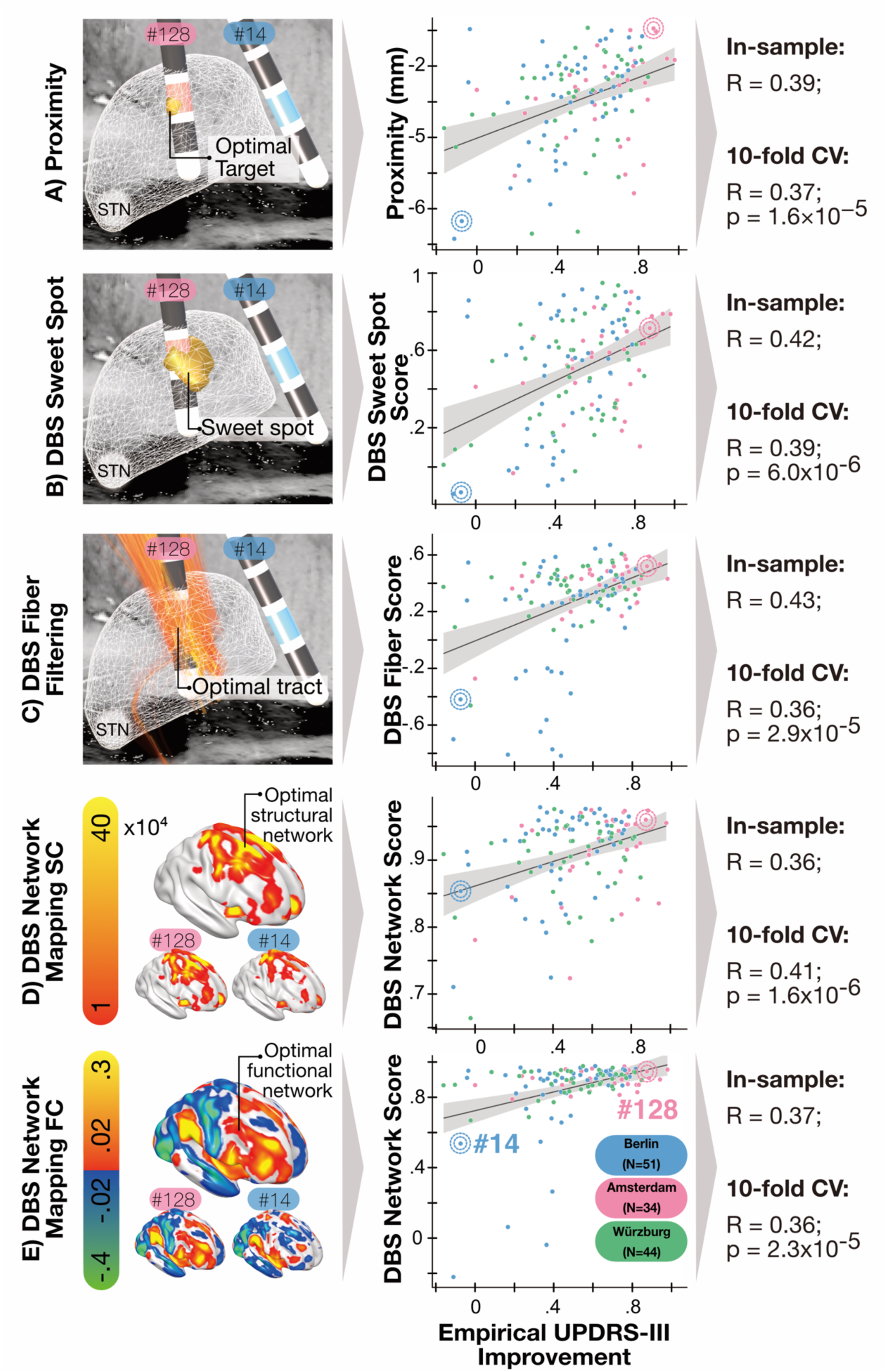
Imaging-based DBS models and their association with clinical outcomes. Five distinct methods were used to generate models that associated clinical improvements with the stimulation site. The validity of these models was examined in a circular manner (in-sample) and via 10-fold cross-validation (CV). P-values are not reported for in-sample calculations due to the circular nature of these results. The left column of the figure depicts a 3D visualization of the model. The golden color represents regions associated with the clinical improvement. The positions of electrodes with labeled stimulated contacts (A-C) along with network profiles (D-E) are shown for two example patients (patient #14 with low empirical improvement and #128 with high empirical improvement). The middle column shows the correlation between the model-based scores and the actual empirical improvements. The positions of the two example patients in the correlation plots are indicated by circles of corresponding colors. The grey shaded area in the plot represents a 95% confidence interval. A) Patients with electrodes closer to the optimal stimulation coordinate (Proximity), as calculated by the Euclidean distance, had higher relative clinical improvement. B) Patients whose E-fields peaked in regions overlapping with the sweet spot received higher scores (DBS Sweet Spot Scores), while those with E-fields that peaked elsewhere tended to receive lower scores. C) Patients whose fiber profile correlated with the modeled optimal fiber profile (DBS Fiber Score) showed on average higher empirical clinical improvements. D) Structural network maps associated with clinical improvement showed stronger correlations with connectivity profiles (DBS Network Score) in patients with higher empirical clinical improvements. E) Optimal functional connectivity profiles more closely resembled the DBS-related patient network profiles (DBS Network Score) in patients with higher empirical clinical improvement. STN – subthalamic nucleus, CV – cross-validation, SC – structural connectivity, FC – functional connectivity, DBS – deep brain stimulation.

### Predicting outcomes in unseen heterogeneous group-level data

The joint model explained 23% of variance in clinical improvement within the discovery cohort (in-sample: *R* = 0.48, *R^2^* = 0.23) and 16 % using ten-fold cross-validation (*R* = 0.40, *R^2^* = 0.16, *p* < 0.001) (Figure 3A). The predictors influenced the clinical scores in the following order: DBS Fiber Filtering score, DBS Sweet Spot score, DBS Network Mapping SC score, Proximity, and DBS Network Mapping FC score (Figure 3B, stability of *ß* estimates is shown in Supplementary Methods 3, Supplementary Figure 3). In the validation dataset, the model explained 11 % of variance in clinical improvement (*R* = 0.34, *R^2^* = 0.11, *p* = 0.001) (Figure 3C). See results for individual methods in the Supplementary material (Supplementary Results 2). Importantly, the variance was not obtained by squaring the *R*-value, but by computing the coefficient of determination, in line with the community standards proposed to demonstrate evidence of “prediction.”^26^ Alternatively, squaring the *R*-value yielded a comparable *R^2^*(*R* = 0.34: *R^2^* = 0.12 at *p* = 0.001). This value could be interpreted as the amount of variance explained in ranks of outcomes across the unseen cohort. While all modalities contributed to the ridge regression, fiber filtering and sweet spot mapping had the strongest impact on the model performance.

**Figure 3.**
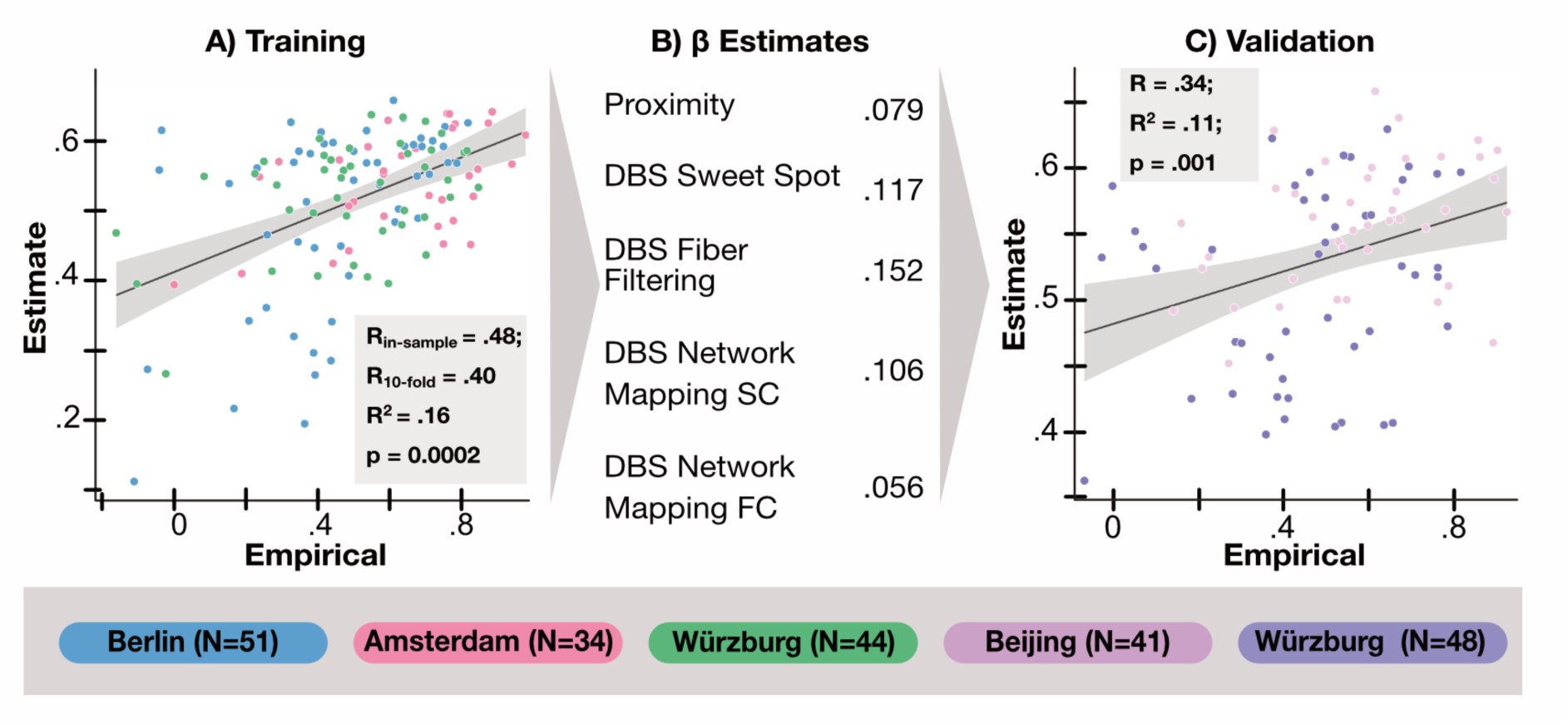
Validation of a combined model of optimal deep brain stimulation (DBS) profile. Our combined model, trained on the discovery dataset, was used to estimate clinical improvements in unseen patients. The model was fitted using the ridge regression method with a *λ* parameter of 50 (Supplementary methods 2, Supplementary Figure 2). The grey shaded area in the plot represents a 95% confidence interval. A) The optimal stimulation models, determined by a data-driven method, significantly correlated with the empirical clinical improvements both in in-sample and in a ten-fold cross-validation design. B) The strongest predictor of clinical improvement was the fiber filtering score (*ß* = 0.152), followed by sweet-spot score (*ß* = 0.117), structural connectivity network mapping score (*ß* = 0.106), proximity to an optimal coordinate (*ß* = 0.079), and functional connectivity network mapping score (*ß* = 0.056). C) This model’s estimates explained 11% of the variance in an out-of-sample validation.

### Matching optimal electrode contacts in prospectively acquired patients

The model explained 11 % of variance in the retrospective, heterogeneous (“noisy”) hold-out test cohort, matching our literature-informed theoretical expectations as well as the approximate explained variance in smaller studies with comparable aims. ^1,9,10,13,27^ However, the key question of whether our model could effectively inform DBS programming in individual patients remained open. To test this, we analyzed a prospectively collected dataset in which each of eight contacts on 21 segmented DBS electrodes (BSci Vercise Directed) was stimulated at a fixed amplitude of 2 mA. Clinical responses were recorded after overnight withdrawal of dopaminergic medication.^28^ Contralateral clinical responses for each electrode were first analyzed separately by i) correlating the model estimates with clinical improvements across the eight contacts and ii) comparing the contact with the highest estimated score with at least one of the best clinical contacts (Figure 4 and 5A). The average correlation value across electrodes was 0.52 ± 0.27 (range 0.01 – 0.92) (Figure 5B). All correlations were positive. High correlations (*R* > 0.7) were present in seven cases. While in some cases correlations were weaker (e.g. *R* = 0.12, *p* = 0.770; Figure 4; E17), the model still correctly identified the optimal clinical contact. Conversely, in other instances, a comparatively high correlation of *R* = 0.65, *p* = 0.080 (Figure 4; E10) did not correctly identify the optimal contact. We must emphasize that these data involved directional leads, where a stimulation level refers to the ring of the lead. Identifying the optimal stimulation level would already prove clinically relevant by reducing programming time.^6^ Our model achieved this success in 71% of cases (chance level 32%, *p_binomial_* = 0.0003). The optimal contact was correctly selected as the top one 38% of the time (chance level 18%, *p_binomial_* = 0.0278), selected in the top two contacts 57% of the time (chance level 35%, *p_binomial_* = 0.0281), and in the top three contacts 81% of the time (chance level 48%; *p_binomial_* = 0.0025). Finally, contacts associated with the lowest empirical improvement were ranked lower than those with highest empirical improvement in 90% of cases. Figure 5A shows details of estimated and empirical contacts for all cases. Results of individual methods are in the Supplementary material (Supplementary Results 2, Supplementary Figure 6 and Supplementary Table 5). As can be visually appreciated, the model suggestions were neighboring or matching the empirical suggestions in all but one electrode. In this electrode (E21), the suggested contact was not adjacent to the best clinical contact but was itself the second-best clinical contact. The linear mixed-effects model (LME), controlling for the patient as the random effect component, shows a moderate to strong positive correlation (*R* = 0.65, *p* < 0.001) between the estimated and empirical improvements. However, the model tends to underestimate the empirical improvements. With every 1-unit increase in the estimated improvement, the empirical improvement increased on average by 0.43 units (*p* < 0.001) (Figure 5C). The model was also able to capture the variance in both akinesia and tremor symptoms (Supplementary Figure 7).

**Figure 4.**
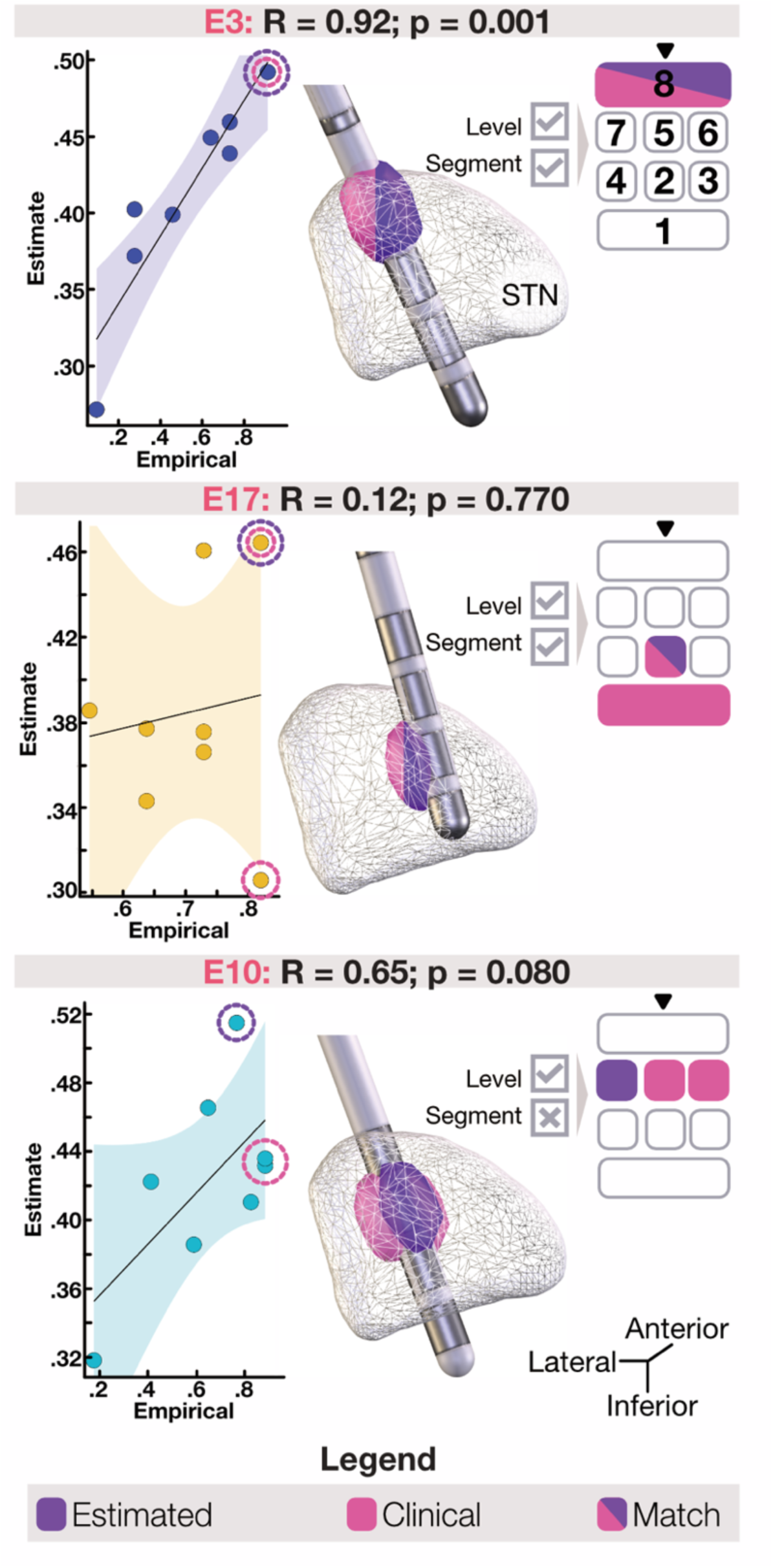
Estimates of clinical improvement in patients with Parkinson’s disease in the contact-wise stimulation cohort. The combined model was used to estimate clinical improvements for 21 electrodes based on contact-wise stimulation data. Each contact had been assigned an empirical clinical improvement by the clinician. Three example patients are shown. For each patient, we show correlation plots between the model-estimated clinical improvement for each contact on the electrode and the empirical clinical improvement assigned by the neurologist (left side of the figure). In the center, we show the electrode’s position within the subthalamic nucleus (STN; white mesh) and stimulation volumes from the model-selected (purple) and empirically selected (pink) optimal contacts. A schematic visualization is shown on the right side. Downward-facing black triangles indicate the location of the electrode marker. In two of the three patients, clinicians assigned identical empirical clinical improvement scores to two contacts. Scores were still assigned individually to each contact during the monopolar review process. For patients E3 and E17, the estimated clinical contact matched the empirical one. For patient E10, the estimated contact did not match the empirically selected one but matched the level at which the contact was located.

**Figure 5.**
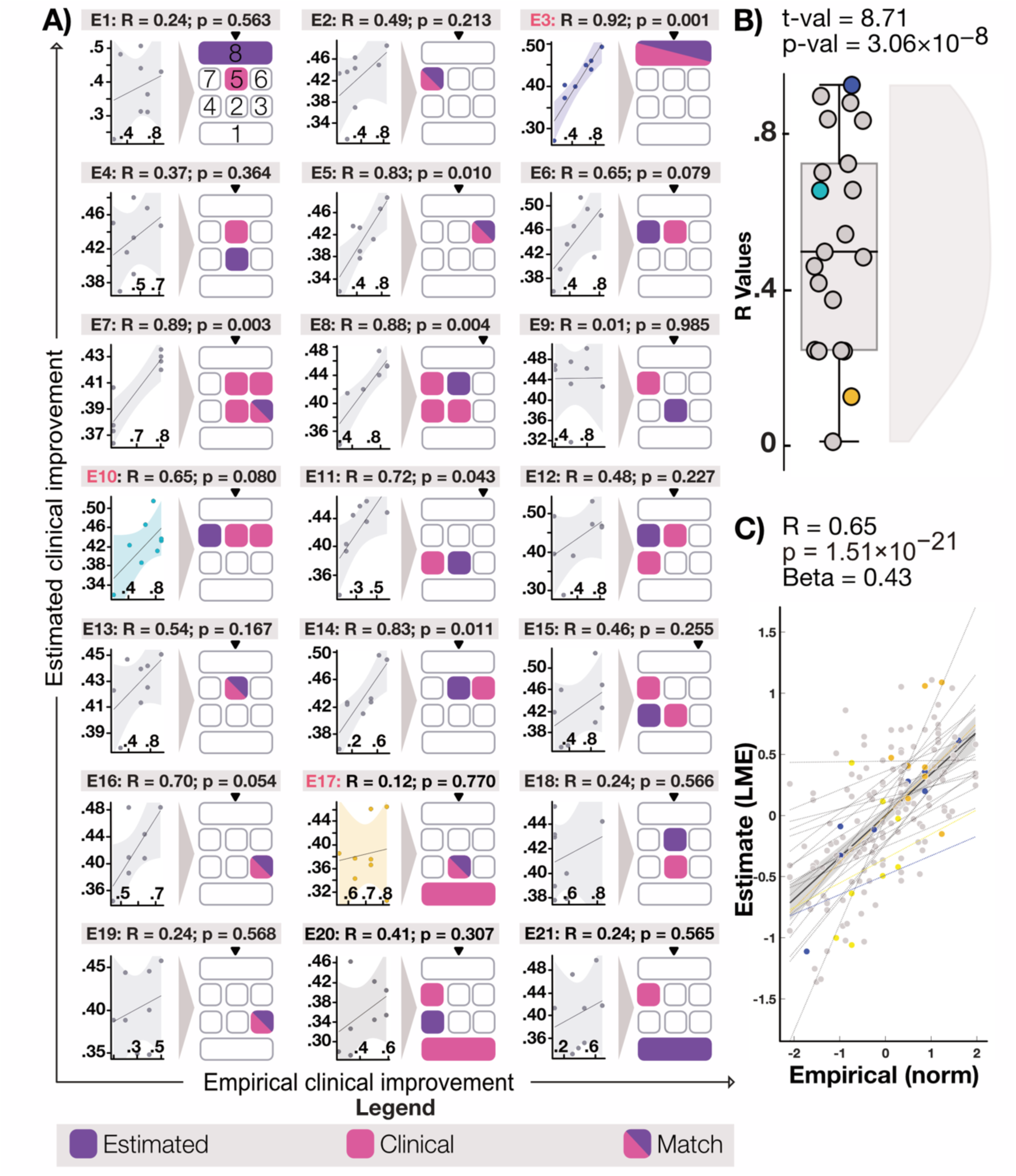
Estimates of clinical improvement for each electrode in the contact-wise stimulation cohort. The combined model estimated clinical improvements for individual contacts of 21 electrodes (labeled as E1 – E21) based on their specific placements. A) In the correlation plots, each point represents an electrode contact, with the x-axis showing its empirical clinical improvement and the y-axis displaying the estimated improvement. Color-coded plots (E3, E10, E17) correspond to the electrodes shown in more detail in Figure 4. To the right of the correlation plots, each electrode is shown with its eight contacts (1 to 8), indicating whether it was identified as the contact with the highest clinical improvement (empirically chosen: pink; chosen by our model: purple). Downward-facing black triangles indicate the location of the electrode marker. B) Distribution of correlation values between model estimates and empirical clinical improvements. C) Results of a linear mixed-effects model, with intercept adjusted for each electrode.

## Discussion

Image guidance is a promising approach to maximize the efficacy of DBS. However, the optimal imaging-based strategy for guiding DBS programming remains unclear, with numerous approaches proposed in the scientific literature.^5,9,29^ While the concept seems like a natural choice, good evidence for clear clinical utility is lacking.^30^ To address this gap, we leveraged multiple state-of-the-art concepts to analyze the effects of an exceptionally large cohort of 604 subthalamic stimulation sites to create and validate DBS models associated with clinical benefit in PD. Both prior literature and the present empirical results suggested that image guidance models could be expected to account for approximately 10% of variance in clinical improvements in an unseen heterogeneous group-level retrospective STN-DBS cohort of PD patients. Importantly, this held true even for our advanced multi-scale model, which combined imaging predictors from five neuroimaging modalities using highly sophisticated methods. However, when controlling for numerous nuisance factors (sources of “noise”) present in typical group-level data, the same model was able to suggest correct or neighboring DBS contacts in individual patients in all cases.

These results suggest clear relevance of image guidance for clinical practice when used to identify optimal contacts in individual patients. The model matched the clinician’s choice in determining the optimal stimulation level in 71% of cases. Identifying the correct stimulation level alone could reduce the programming time by 75% for conventional stimulation and by 62.5% for directional stimulation. This is particularly remarkable for segmented electrode designs, where contact distances are comparable to imaging resolution, even when models were originally trained on heterogeneous group-level data. This heterogeneity in the training data might be considered a strength rather than a limitation, potentially contributing to the model’s success in suggesting optimal contacts in our prospectively acquired patients. Evidence from other fields suggests that identifying brain stimulation targets based on heterogeneous data from different modalities will prevent overfitting and lead to robust models that will more likely generalize to novel data.^31^ This matches our results, where circular and cross-validated correlations were similar, indicating a low risk of overfitting, and the similar amount of variance explained in the “noisy” training and test datasets. Moreover, the successful identification of the optimal stimulation contact suggests that these models trained to explain variance between-subjects in fact explain more meaningful variance within-subjects. Instead, when using the same models to predict clinical outcomes following DBS in ‘noisy’ cohorts, electrode placement remains key but other factors seem at least equally important.

Our results confirm optimal target sites for STN-DBS on a coordinate, sweet-spot, tract and network level and are directly relevant for clinical practice. Both the optimal coordinate and sweet-spot of our model pointed to the Bejjani line,^32^ widely used for surgical targeting in clinical practice. Moreover, the optimal coordinates and sweet-spots precisely matched multiple published reports.^33,34^ At the tract and network levels, our identified connections are in agreement with most historical and modern previous reports.^35,36^ While confirmatory to published reports, given the large cohort of patients, our result may represent a “mature” model of optimal stimulation targets on five levels of analysis whose performance has now been properly quantified and used to suggest stimulation contacts with a high success rate in unseen data. These results lay the groundwork for applying our model in prospective clinical trials that could ultimately save time for both clinicians and patients, making DBS programming more efficient, accurate and deliberate.

Our study is subject to multiple limitations. First, the model was created based on retrospective heterogeneous data from multiple institutions. These cohorts differed in terms of post-surgical data collection times, clinical centers, electrode models, neurosurgeons, neurologists and countries. However, as argued above, this choice was deliberate to maximize robustness and prevent overfitting to a single surgeon or center.^31^ Second, none of the datasets analyzed contained information about side effects. This information could certainly increase the clinical utility of our model, since for now, it is making decisions only based on motor effects. Third, for this line of work, nonlinear registrations between native patient and template space are required, which, despite decade-long improvements of the underlying methodology, will always introduce a certain amount of bias. To minimize this bias, we used a state-of-the-art DBS imaging pipeline designed and optimized for DBS imaging analyses.^1^ Last, biophysical modeling and sweet-spot statistics may be carried out in various ways, and while our study tested several methodological concepts, it would not be possible to compare every single method that has been proposed over the years.^37^

In conclusion, we introduce a new multi-level DBS image guidance model which explained a significant amount of variance in two validation datasets and suggested clinically sensible electrode contact choices. By computationally estimating the clinical impact of each contact within an individual patient, it may potentially guide clinicians toward the most optimal contacts for initial DBS programming, reducing both the duration and burden of programming and improving clinical practice.

## Data Availability

All code used to analyze the dataset is openly available within Lead-DBS (including sweet
spot mapping, fiber filtering and network mapping software:
https://github.com/leaddbs/leaddbs).
Cohort-wise demographic and clinical outcomes from the training and the test cohorts are
available in Supplementary Table 2 and 3. We cannot openly share patient imaging data due
to data sharing and privacy regulations, but they can be made available upon request to the
corresponding primary investigator. The corresponding author and the principal investigator (PZ and AH) commit to returning data requests within a time frame of 30 days.

## Acknowledgements

The authors would also like to thank Dr. Simón Oxenford, developer of the WarpDrive tool, which was used to enhance the data normalization process. Furthermore, we thank the patients and their families for their participation in the clinical trials that allowed the collection of data for this research.

## Author Contributions

P.Z., N.R., and A.H. contributed to conceptualization of the study, study design, and research direction. N.R., and A.H. contributed to supervision. C.V.L., T.B, J.N.P.-S, T.A.D, M.T.B, V.V.-V., M.R, J.V, V.J.J.O, R.M.A.B, X.X., Z.L., C.Y., A.A.K, contributed to data collection and data organization. P.Z., N.R., A.H., N.L., K.B., G.M.M, A.R.P contributed to data analysis and methodology. P.Z., A.H., N.R., G.M., C.V.L., T.B, J.N.P.-S, T.A.D contributed to electrode localization. I.A.S, L.L.G, B.H.B, B.H., contributed to critical revision of the theoretical background and manuscript. A.H., N.R., G.M.M, K.B., M.T.B, M.D.F., J.N.P.-S, C.V.L., T.A.D., S.R.S, K.R. contributed to writing – review & editing. A.H., N.R., M.D.F, G.M.M discussed the results. P.Z contributed to writing – original draft & editing.

## Funding

Scholarship from the Einstein Center for Neurosciences Berlin (PZ, BH, and IAS)

Deutsche Forschungsgemeinschaft, DFG, Project Number 502436811 (JNP-S)

Schilling Foundation (AH)

Deutsche Forschungsgemeinschaft, 424778381 – TRR 295 (AH)

Deutsches Zentrum für Luft- und Raumfahrt (DynaSti grant within the EU Joint Programme Neurodegenerative Disease Research, JPND) (AH)

National Institutes of Health (R01MH130666, 1R01NS127892-01, 2R01 MH113929 & UM1NS132358) (AH)

New Venture Fund (FFOR Seed Grant) (AH)

## References

1. Neudorfer C, Butenko K, Oxenford S, et al. Lead-DBS v3.0: Mapping deep brain stimulation effects to local anatomy and global networks. NeuroImage. 2023;268:119862. doi:10.1016/j.neuroimage.2023.119862

2. Lozano AM, Lipsman N, Bergman H, et al. Deep brain stimulation: current challenges and future directions. Nat Rev Neurol. 2019;15(3):148–160. doi:10.1038/s41582-018-0128-2

3. Behnke JK, Peach RL, Habets JGV, et al. Long-Term Stability of Spatial Distribution and Peak Dynamics of Subthalamic Beta Power in Parkinson’s Disease Patients. Movement Disorders. 2025;n/a. doi:10.1002/mds.30169

4. Busch JL, Kaplan J, Bahners BH, et al. Local Field Potentials Predict Motor Performance in Deep Brain Stimulation for Parkinson’s Disease. Movement Disorders. 2023;38(12):2185–2196. doi:10.1002/mds.29626

5. Roediger J, Dembek TA, Wenzel G, Butenko K, Kühn AA, Horn A. StimFit—A Data-Driven Algorithm for Automated Deep Brain Stimulation Programming. Movement Disorders. 2022;37(3):574–584. doi:10.1002/mds.28878

6. Roediger J, Dembek TA, Achtzehn J, et al. Automated deep brain stimulation programming based on electrode location: a randomised, crossover trial using a data-driven algorithm. The Lancet Digital Health. 2023;5(2):e59–e70. doi:10.1016/S2589-7500(22)00214-X

7. Miocinovic S, Noecker AM, Maks CB, Butson CR, McIntyre CC. Cicerone: stereotactic neurophysiological recording and deep brain stimulation electrode placement software system. Acta Neurochir Suppl. 2007;97(Pt 2):561–567.

8. Maks CB, Butson CR, Walter BL, Vitek JL, McIntyre CC. Deep brain stimulation activation volumes and their association with neurophysiological mapping and therapeutic outcomes. J Neurol Neurosurg Psychiatr. 2009;80(6):659–666. doi:10.1136/jnnp.2007.126219

9. Rajamani N, Friedrich H, Butenko K, et al. Deep brain stimulation of symptom-specific networks in Parkinson’s disease. Nat Commun. 2024;15(1):4662. doi:10.1038/s41467-024-48731-1

10. Hollunder B, Ostrem JL, Sahin IA, et al. Mapping dysfunctional circuits in the frontal cortex using deep brain stimulation. Nat Neurosci. 2024;27(3):573–586. doi:10.1038/s41593-024-01570-1

11. Horn A, Reich M, Vorwerk J, et al. Connectivity Predicts deep brain stimulation outcome in Parkinson disease. Ann Neurol. 2017;82(1):67–78. doi:10.1002/ana.24974

12. Li N, Baldermann JC, Kibleur A, et al. A unified connectomic target for deep brain stimulation in obsessive-compulsive disorder. Nature Communications. 2020;11(1):3364. doi:10.1038/s41467-020-16734-3

13. Ríos AS, Oxenford S, Neudorfer C, et al. Optimal deep brain stimulation sites and networks for stimulation of the fornix in Alzheimer’s disease. Nat Commun. 2022;13(1):7707. doi:10.1038/s41467-022-34510-3

14. Reich MM, Horn A, Lange F, et al. Probabilistic mapping of antidystonic effect of pallidal neurostimulation: multicentre imaging study. Brain. Published online January 12, 2019:1–80.

15. Meyer GM, Hollunder B, Li N, et al. Deep Brain Stimulation for Obsessive-Compulsive Disorder: Optimal Stimulation Sites. Biological Psychiatry. Published online December 2023:S0006322323017857. doi:10.1016/j.biopsych.2023.12.010

16. Alonso F, Zsigmond P, Wårdell K. Influence of Virchow-Robin spaces on the electric field distribution in subthalamic nucleus deep brain stimulation. Clinical Neurology and Neurosurgery. 2021;204:106596. doi:10.1016/j.clineuro.2021.106596

17. Avants BB, Tustison NJ, Song G, Cook PA, Klein A, Gee JC. A Reproducible Evaluation of ANTs Similarity Metric Performance in Brain Image Registration. Neuroimage. 2011;54(3):2033–2044. doi:10.1016/j.neuroimage.2010.09.025

18. Fonov V, Evans AC, Botteron K, Almli CR, McKinstry RC, Collins DL. Unbiased average age-appropriate atlases for pediatric studies. NeuroImage. 2011;54(1):313–327. doi:10.1016/j.neuroimage.2010.07.033

19. Oxenford S, Ríos AS, Hollunder B, et al. WarpDrive: Improving spatial normalization using manual refinements. Medical Image Analysis. 2024;91:103041. doi:10.1016/j.media.2023.103041

20. Horn A, Li N, Dembek TA, et al. Lead-DBS v2: Towards a comprehensive pipeline for deep brain stimulation imaging. NeuroImage. 2019;184:293–316. doi:10.1016/j.neuroimage.2018.08.068

21. Husch A, V. Petersen M, Gemmar P, Goncalves J, Hertel F. PaCER - A fully automated method for electrode trajectory and contact reconstruction in deep brain stimulation. Neuroimage Clin. 2017;17:80–89. doi:10.1016/j.nicl.2017.10.004

22. Horn A, Kühn AA. Lead-DBS: A toolbox for deep brain stimulation electrode localizations and visualizations. NeuroImage. 2015;107:127–135. doi:10.1016/j.neuroimage.2014.12.002

23. Dembek TA, Hoevels M, Hellerbach A, et al. Directional DBS leads show large deviations from their intended implantation orientation. Parkinsonism & Related Disorders. 2019;67:117–121. doi:10.1016/j.parkreldis.2019.08.017

24. Butenko K, Bahls C, Schröder M, Köhling R, van Rienen U. OSS-DBS: Open-source simulation platform for deep brain stimulation with a comprehensive automated modeling. PLoS Comput Biol. 2020;16(7):e1008023. doi:10.1371/journal.pcbi.1008023

25. Baldermann JC, Melzer C, Zapf A, et al. Connectivity Profile Predictive of Effective Deep Brain Stimulation in Obsessive-Compulsive Disorder. Biol Psychiatry. 2019;85(9):735–743. doi:10.1016/j.biopsych.2018.12.019

26. Poldrack RA, Huckins G, Varoquaux G. Establishment of Best Practices for Evidence for Prediction: A Review. JAMA Psychiatry. 2020;77(5):534–540. doi:10.1001/jamapsychiatry.2019.3671

27. Sobesky L, Goede L, Odekerken VJJ, et al. Subthalamic and pallidal deep brain stimulation: are we modulating the same network? Brain. 2022;145(1):251–262. doi:10.1093/brain/awab258

28. van der Linden C, Berger T, Brandt GA, et al. Accelerometric Classification of Resting and Postural Tremor Amplitude. Sensors (Basel). 2023;23(20):8621. doi:10.3390/s23208621

29. Horn A. The impact of modern-day neuroimaging on the field of deep brain stimulation. Current Opinion in Neurology. 2019;32(4):511–520. doi:10.1097/WCO.0000000000000679

30. Hines K, Noecker AM, Frankemolle-Gilbert AM, et al. Prospective Connectomic-Based Deep Brain Stimulation Programming for Parkinson’s Disease. Movement Disorders. Published online October 21, 2024:mds.30026. doi:10.1002/mds.30026

31. Siddiqi SH, Kording KP, Parvizi J, Fox MD. Causal mapping of human brain function. Nat Rev Neurosci. 2022;23(6):361–375. doi:10.1038/s41583-022-00583-8

32. Bejjani BP, Dormont D, Pidoux B, et al. Bilateral subthalamic stimulation for Parkinson’s disease by using three-dimensional stereotactic magnetic resonance imaging and electrophysiological guidance. Journal of Neurosurgery. 2000;92(4):615–625. doi:10.3171/jns.2000.92.4.0615

33. Akram H, Sotiropoulos SN, Jbabdi S, et al. Subthalamic deep brain stimulation sweet spots and hyperdirect cortical connectivity in Parkinson’s disease. NeuroImage. 2017;158:332–345. doi:10.1016/j.neuroimage.2017.07.012

34. Bot M, Schuurman PR, Odekerken VJJ, et al. Deep brain stimulation for Parkinson’s disease: defining the optimal location within the subthalamic nucleus. J Neurol Neurosurg Psychiatr. Published online January 20, 2018:jnnp-2017-316907-7. doi:10.1136/jnnp-2017-316907

35. Treu S, Strange B, Oxenford S, et al. Deep brain stimulation: Imaging on a group level. NeuroImage. 2020;219:117018. doi:10.1016/j.neuroimage.2020.117018

36. Hassler R, Riechert T, Mundinger F, Umbach W, Ganglberger JA. PHYSIOLOGICAL OBSERVATIONS IN STEREOTAXIC OPERATIONS IN EXTRAPYRAMIDAL MOTOR DISTURBANCES. Brain. 1960;83(2):337–350. doi:10.1093/brain/83.2.337

37. Dembek TA, Baldermann JC, Petry-Schmelzer JN, et al. Sweetspot Mapping in Deep Brain Stimulation: Strengths and Limitations of Current Approaches. Neuromodulation: Technology at the Neural Interface. 2022;25(6):877–887. doi:10.1111/ner.13356

