## Supplementary File for "Multimodal Image Guidance in Subthalamic Deep Brain Stimulation for Parkinson’s Disease"

### Supplementary Text 1

### Factors contributing to clinical improvements in STN-DBS for Parkinson's disease

Before developing models aimed at (1) explaining clinical variance in group level data and (2) suggesting optimal contacts in individual patients, we began with theoretical considerations and literature review to inform expectations for both tasks. We searched PubMed and Google Scholar databases for studies that explored factors, beyond deep brain stimulation electrode placement (DBS), which would influence clinical improvement in Parkinson's disease (PD) after DBS surgery. Most studies did not report concrete values for the variance explained by specific factors. For each article, we therefore estimated the expected contributions, based on the values reported. In most cases, we were rather careful not to inflate the influence of specific factors, i.e. we leaned toward the lower end of reported values.

One of the most comprehensive studies was conducted by Cavallieri et al.[1], who analyzed predictors of long-term outcomes in patients with PD who underwent DBS in a long-term cohort of 138 patients and short-term cohort of 357 patients. The study assessed the contribution of ten different predictors: Age at the surgery, onset age, young onset Parkinson's disease (YOPD), presence of white matter hyperintensities on brain MRI, Mattis Dementia Rating Scale (MDRS), Frontal Score, part III of the Movement Disorders Society Unified Parkinson's Disease Rating Scale (MDS-UPDRS) off-medication before the surgery, Hoehn & Yahr stage off-medication, levodopa responsiveness and tremor dominant phenotype (TDP). Apart from TDP, all factors contributed significantly to the short-term cohort. In the long-term cohort, significant contributors were frontal lobe dysfunction, disease severity, and the presence of vascular changes. Since the variance explained by these predictors (and their covariance structure) was not reported, we estimated the contributions based on the reported standardized ß-coefficients (Reference 5 in Figure 1A and the Supplementary Table 1). Other studies that we reported in a category termed "disease state" focused on the contribution of anxiety and depression (Reference 11 in Figure 1A and Supplementary Table 1).[2] This study reported the variance explained in DBS-related improvement, which we rounded down to avoid inflated effects. The idea that emotional well-being affects DBS outcomes is not new, but most evidence remains anecdotal.[3]

Beyond disease state, anatomical abnormalities, especially near the target nucleus, can influence clinical response to DBS. For example, microstructural changes such as the presence of Virchow-Robin spaces near the electrode influence the spread of electric field beyond the target nucleus (Reference 9 in Figure 1A and Supplementary Table 1).[4] Neither image guidance models nor most biophysical modeling approaches account for these structural abnormalities in individual patients. As a result, predicting clinical outcomes for individual contacts in such patients may yield imprecise results. Besides microstructural abnormalities, cortical atrophy present in paracentral areas and superior frontal cortex may lead to decreased responsiveness of DBS (Reference 10 Figure 1A and Supplementary Table 1).[5] If we enter two patients with an ideal (and equal) electrode placement within the STN into our model, the model would predict the same optimal improvement for both. However, if one of these patients had substantial atrophy in frontal areas, clinical effects would likely be less optimal in this patient compared to the other one.

Although the UPDRS-III is well established and healthcare providers are well trained to assess the motor symptoms of PD, there is always an expected error between different clinical assessments (be it the intra- or the inter-rater error). Studies that looked into this issue[6–9] reported excellent reliability based on non-significant statistical group differences. However, small margins of error were observed. For these reasons, we did not exclude the small variance explained by these two factors, but we decided to keep it rather low at 2% for both intra- and inter-rater reliability, respectively. It is important to note that our clinical outcomes were assessed across various DBS centers, countries and continents (References 1-4 in Figure 1A and Supplementary Table 1).

Since our analysis focused on image guidance, model performance depends on how imaging data was collected, processed and analyzed. Estimating the variance explained by our selection of neuroimaging methods is not straightforward, since we use a combination of different methods that have not been compared head-to-head. Further, various methods have been proposed to approximate stimulation effects[10,11] (references 5 and 6 in Figure 1A and the Supplementary Table 1). In our methods, we make assumptions about the physical properties of the electric field imposed by the electrode, the conductivity of axons, as well as the intactness of the brain. Moreover, inaccuracies in electrode reconstruction in both the patient's native space and standard stereotactic space may increase modeling uncertainty (reference 7 in Figure 1A and the Supplementary Table 1). Rajamani et al.[12] examined how adding uncertainty (jitter) to their electrode location affected model performance based on optimal stimulation tracts. Although the exact amount of variance introduced was not reported, the average spatial correlation between their baseline model and 1,000 jittered models was .80. Based on these assumptions, we estimated approximately 10% of variance explained to be attributed to the neuroimaging factor. We visualize all aforementioned factors in Figure 1A.

While the earlier considerations addressed explaining variance in clinical improvement using group-level data, the nuisance variables differ substantially when the goal is to suggest optimal contacts for individual patients. The reason is that here, most of the abovementioned variables would be fixed, i.e. would not contribute noise within the individual patient. For instance, the patient will have the same age, PD subtype, comorbidities, etc. when our question is which of their electrode contacts would perform the best. We still identified three categories that would represent nuisance variables in individual patients, which are inaccuracy introduced by imaging, microstructural anatomical changes (which may have differential effects on individual electrode contacts) and a high covariance structure in clinical effects across neighboring contacts.

Imaging (References 6-8 in Figure 1B and Supplementary Table 1)[10–12] and anatomical factors (Reference 9 in Figure 1B and Supplementary Table 1)[4] were already discussed previously (Figure 1A). As we are still analyzing neuroimaging data, the processing pipeline may introduce uncertainty, and anatomical deviations could further affect the model's accuracy. Microstructural anatomical changes within the target region such as structural lesions or Virchow-Robin spaces could contribute to unaccounted noise since they may differentially impact the clinical response of certain electrode contacts, but not others. The third factor, clinical measures, addresses that stimulating adjacent electrode contacts will often lead to similar effects, and it may not always be straightforward to identify a single contact that is clearly the best, especially when dealing with segmented electrode designs. Factors such as testing sequence, patient fatigue, and intra-rater variability can make the identification of the "optimal contact" somewhat imprecise and subjective, even in meticulously and prospectively acquired data, as in the present study. In our models, which are based on anatomical positions, no two contacts will ever be assigned identical values. This is because each contact is always located in a unique anatomical position, so the model will assign (slightly) distinct values for each contact within a given patient. With these considerations, we estimate that, while the electrode contact choice could theoretically account for all the variance, image-guided models should optimally only be able to account for around 70% of the variance when suggesting optimal contacts for individual patients. We visualize all these factors in Figure 1B. It is also important to note, that imaging resolution is relatively low and capturing within-contact differences at this level of precision is still quite astonishing (Figure 1C).

Complete list of references from Figure 1: [1-4][6–9], [5][1], [6-8][10–12], [9][4], [10][5], [11][2]

**Supplementary Table 1.** Factors contributing to clinical improvement after deep brain stimulation (reference table).

| Citation number | First Author | Title | Year | Factor | Category | Statistics | | | |
|---|---|---|---|---|---|---|---|---|---|
| | | | | | | R | R^2 | Other | p |
| 1 | **Richards et al.,** | Interrater Reliability of the Unified Parkinson's Disease Rating Scale Motor Examination | 1994 | Intra-/Inter-rater reliability | Neurologist | N/A | N/A | Total motor score ICC (0.82) | N/A |
| 2 | **Bennett et al.,** | Metric properties of nurses' ratings of parkinsonian signs wit ha modified Unified Parkinson's Disease Rating Scale | 1997 | Intra-/Inter-rater reliability | Neurologist | Average correlation for total UPDRS-III (0.92) | N/A | The overall stability of the global parkinsonism measure (0.90) | N/A |
| 3 | **Siderowf et al.,** | Test–Retest Reliability of the Unified Parkinson's Disease Rating Scale in Patients with Early Parkinson's Disease: Results from a Multicenter Clinical Trial | 2002 | Intra-/Inter-rater reliability | Neurologist | N/A | N/A | ICC for the total UPDRS = 0.92 | N/A |
| 4 | **Metman et al.,** | Test-Retest Reliability of UPDRS-III Dyskinesia Scales, and Timed Motor Tests in Patients with Advanced Parkinson's Disease: An Argument Against Multiple Baseline Assessments | 2004 | Intra-/Inter-rater reliability | Neurologist | N/A | N/A | ICC of UPDRSIII off state = 0.90<br>ICC of UPDRSIII on state = 0.89 | N/A |
| 5 | **Cavallieri et al.,** | Predictors of Long-Term Outcome of Subthalamic Stimulation in ParkinsonDisease | 2020 | Age at the surgery | Demographics | N/A | N/A | (-0.186) Standardized ß coefficient | <0.001 |
| | | | | Onset age | Demographics | | | (-0.231) Standardised ß coefficient | <0.001 |
| | | | | YOPD | Disease State | | | (0.213) Standardised ß coefficient | <0.001 |
| | | | | Presence of WMH on brain MRI | Disease State | | | (-0.180) Standardised ß coefficient | 0.002 |
| | | | | MDRS | Disease State | | | (0.167) Standardised ß coefficient | 0.002 |
| | | | | Frontal Score | Disease State | | | (0.272) Standardised ß coefficient | <0.001 |
| | | | | MDS-UPDRS part III off-medication | Disease State | | | (0.320) Standardised ß coefficient | <0.001 |
| | | | | H&Y stage off-medication | Disease State | | | (0.194) Standardized ß coefficient | <0.001 |
| | | | | Levodopa responsivness | Disease State | | | (0.207) Standardised ß coefficient | <0.001 |
| | | | | TD phenotype | Disease State | | | (0.159) Standardised ß coefficient | 0.003 |
| 6 | **Maks et al.,** | Deep brain stimulation activation volumes and their association with neurophysiological mapping and therapeutic outcomes | 2009 | Processing (Connectomic modelling) | Imaging | N/A | N/A | N/A | N/A |
| 7 | **Noecker et al.,** | StimVision v2: Examples and Applications in Subthalamic Deep Brain Stimulation for Parkinson's Disease | 2021 | Processing (stimulation volume) | Imaging | N/A | N/A | N/A | N/A |
| 8 | **Rajamani et al.,** | Deep brain stimulation of symptom-specific networks in Parkinson'sdisease | 2024 | Processing (electrode position) | Imaging | Results based on the jittered electrode location correlate with the original results by 80% ( R = 0.80). | N/A | N/A | N/A |
| 9 | **Alonso et al.,** | Influence of Virchow-Robin spaces on the electric field distribution in subthalamic nucleus deep brain stimulation | 2021 | Microstructural changes | Anatomy | N/A | N/A | N/A | N/A |
| 10 | **Muthuraman et al.,** | Effects of DBS in parkinsonian patients depend on the structural integrity of frontal cortex | 2017 | Cortical Thickness (CT) | Disease State | 0.35 (CT in paracentral area) | 0.1225 | F = 8.6 | 0.005 |
| | | | | | | 0.34 (CT in frontal region) | 0.1156 | F = 8.25 | 0.005 |
| 11 | **Soulas et al.,** | Depression and Coping as Predictors of Change After Deep Brain Stimulation in Parkinson's Disease | 2011 | Baseline outcome | Disease State | N/A | 0.077 | ß (0.082) | N/A |
| | | | | Sex, age at surgery, disease duration, MDRS score | Dmographics | | 0.241 | ß (0.140,0.115,0.422*,-0.170) respectively | |
| | | | | WCC-R = problem-focused | Disease State | | 0.001 | ß (-0.039) | |
| | | | | WCC-R = emotion-focused | Disease State | | 0.001 | ß (-0.046) | |
| | | | | WCC-R = social-focused | Disease State | | 0.066 | ß (0.278) | |
| | | | | STAI (trait) | Disease State | | 0.089* | ß (-0.321) | |
| | | | | BDI-II (trait) | Disease State | | 0.133* | ß (-0.380) | |

*ICC: Intraclass correlation coefficient; WCC-R: Ways of coping revised; H&Y stage: Hoehn and Yahr Scalel; MDS-UPDRS-III: MDS Unified Parkinson's Disease Rating Scale; STAI: State Trait Anxiety Inventory; BDI-II: Beck Depression Inventory.*

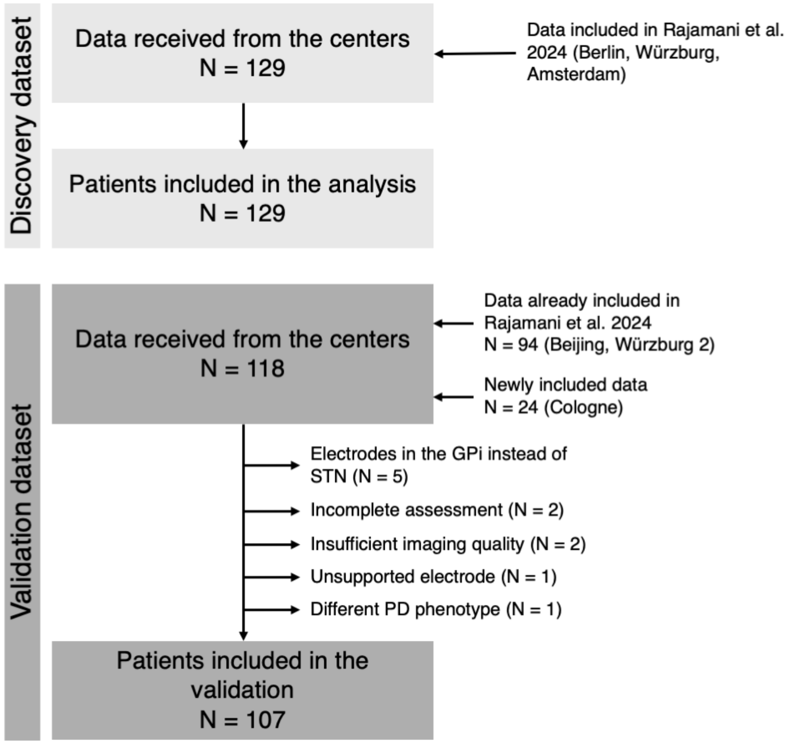

**Supplementary Figure 1: Flowchart of inclusion and exclusion criteria.**

### Supplementary Methods 1
### **Extended DBS modeling methods**

Our modelling approach included four different levels of analysis, using five distinct methods. The first method included the optimal active contact target, expressed as x-, y- and z-coordinates in MNI space (millimeters). After electrode localizations were transformed into a normative space, we extracted the 3-dimensional coordinates of patient-specific contacts selected for chronic stimulation. If multiple contacts per patient were stimulated, the coordinates of these contacts were averaged to obtain a single location per patient. To determine the stimulation coordinates associated with optimal clinical improvements, coordinates were weighted by the $10^{th}$ power of normalized patient-specific relative UPDRS-III improvements. The $10^{th}$ power was chosen to maximize differences between high and low responders, giving greater weight to patients with higher clinical improvement. The resulting coordinates were averaged to yield a single point in 3D space.

Second, we explored the stimulation profile associated with the optimal clinical improvements in a DBS Sweet Spot analysis.[13,14] E-fields estimated for each patient based on their stimulation parameters were thresholded at 100 V/m. Only voxels included in the stimulation volumes of at least 10% of patients were retained. Patient-specific E-field magnitudes were mirrored to the contralateral side as done in previous studies.[12] This was done to increase statistical power, as previous studies found no lateralized effects in the motor UPDRS.[3,13,15,16] Thus, each voxel contained 258 values associated with the E-field magnitudes from the discovery dataset. These values were correlated with clinical improvements, resulting in a profile of voxels associated with optimal (sweet-spot) or suboptimal (sour-spot) clinical improvement. Only significant voxels ($p < .05$) were considered for the model.

Third, we explored the association between fiber tracts and clinical improvements using the DBS Fiber Filtering method.[12,13] The Netstim Atlas V2.2[12] was used to represent fiber tract candidates in the STN region. Like the sweet spot level, we did not expect lateralization of DBS motor effects based on global UPDRS-III scores, therefore streamlines were mirrored across hemispheres to maximize statistical power. To estimate biophysical models, streamlines were warped into each patient's anatomical space. E-field values associated with tract "activations" are typically considered to be in the range of above ~200 V/m.[17] Streamlines falling consistently below this threshold were excluded to avoid spurious correlations without contributing meaningfully to DBS outcomes. To prevent this, we discarded streamlines that were not covered by E-field magnitudes of at least 50 V/m in any patient from the analysis. For similar reasons, we also wanted to discard streamlines that traversed the periphery of electric fields for all patients, i.e., were not clearly modulated by at least a few electrodes. Of the remaining streamlines, only those covered by an E-field magnitude of at least 500 V/m in more than 3 % of all stimulation fields were retained. To determine the optimal streamline profile associated with the maximum DBS related clinical benefit, for each streamline we correlated the patient-specific E-field with the patient-specific clinical outcomes. Positive correlations indicated streamlines associated with greater clinical improvement, while negative correlations indicated less optimal outcomes.

Fourth, we applied the DBS Network Mapping approach using both structural and functional connectomes[3] to estimate cortico-subcortical connectivity, based on voxel-wise correlations. Structural networks were defined using the same atlas as in the fiber-filtering step.[12] Functional networks were defined using resting-state functional MRI (rs-fMRI) from 1,087 scans in the Human Connectome Project[18] stored in a matrix format. To estimate the structural DBS connectivity profile seeding from each patient's stimulation volumes, we calculated the overlap between voxels with the E-fields information and the voxels with the fibers of the normative structural connectome. To estimate the functional connectivity, we first thresholded and binarized the E-fields at a typical value of 200 V/m.[17] Functional connectivity profiles for each patient were then calculated as described in Goede et al.[19] In short, we used a precalculated whole brain voxel-wise connectome based on 1,000 resting-state fMRI scans (four runs each) acquired within the Human Connectome Project[18]. This average voxel-wise connectome is stored as a large adjacency matrix in 2 mm isotropic resolution (i.e. storing the connectivity of each voxel in MNI space). To obtain functional connectivity profiles seeding from each patient's stimulation volumes, the rows of the connectome corresponding were averaged to obtain a whole-brain profile. This procedure was repeated for each patient to acquire their unique functional connectivity fingerprint.

Optimal network profiles for both structural and functional data were calculated using the combined map (C-map) approach, which has shown robust performance in previous work.[3] These maps represent the connectivity profile of stimulation volumes, but are positively associated with clinical outcomes. Namely, first, fingerprint maps were correlated with clinical improvement using Pearson's correlations, resulting in a correlation map (R-map). Next, weighted average maps were calculated by averaging all fingerprints weighted by their respective clinical improvements (A-map). Finally, the R-maps and the A-maps were combined by retaining the A-map voxels with the same sign as the corresponding R-map, resulting in the final C-map.[3]

### Supplementary Methods 2
### **Surrogate variable calculation**

At the E-field level, spatial similarity between each E-field and the sweet-spot model was calculated using Pearson's correlation as done in previous studies.[13,20,21] Here, the surrogate for the clinical improvement was expressed as the spatial correlation coefficient between sweet spots and electric fields. The intuitive interpretation of this is as follows: if a patient's E-field profile spatially resembles the landscape defined as "optimal" by the sweet spot, the model would expect high clinical improvement. Ten-fold cross-validation was carried out analogously for the coordinate, E-field, tract, and network levels.

At the tract level, the surrogate was expressed similarly to the sweet spot model, i.e., by measuring the overlap between E-field magnitudes of a given patient with the optimal streamline model. The same interpretation applies: if the shape of an electric field precisely matched the one defined by the optimal tract landscape, the model would estimate higher DBS response. After performing both in-sample evaluation and ten-fold cross-validation, we correlated the surrogate with the empirical clinical improvement, expecting consistently positive correlation results.

Finally, on a network level, the surrogate was defined using the same metric: each patient's network profile (connectivity fingerprint) was spatially correlated with the optimal stimulation network profile. If the spatial definition of the network profile seeding from the patient's E-fields matched the optimal DBS network profile, the model would estimate optimal DBS response, resulting in higher surrogate values. Again, we applied both in-sample evaluation and ten-fold cross-validations.

Across these models (coordinates, E-fields, tracts, structural and functional networks), each patient of the discovery dataset was assigned five surrogate values (essentially representing similarities between each patient and the model) determined from the in-sample design. These values were later used to fit a ridge regression model, which provided beta estimates for the out-of-sample validation in the hold-out cohort.

### Supplementary Methods 3
### **Ridge regression regularization parameter calculation**

To determine a suitable ridge regression regularization parameter (λ factor), we followed guidelines provided by MathWorks (https://www.mathworks.com/help/stats/ridge.html), where the λ factor is referred to as the variable *k*. Critically, this process was carried out exclusively using the discovery cohort, before any attempts were made to explain variance in the hold-out test dataset. Following the recommended guidelines, we first tested how different *k* values affected coefficient estimates. To do so, we created a matrix of predictors, including the interaction and polynomial terms. We then plotted the standardized coefficients obtained during in-sample cross-validation against the *k* parameter. The goal was for the model to stabilize, i.e., for the standardized coefficients to converge and for the plotted lines to flatten. We tested three different *k* coefficients ($k$ = 10, 50, and 90) on our model from the discovery dataset to assess changes in the correlation between estimated and empirical clinical improvement, as well as changes in beta estimates (Supplementary Figure 2A). The resulting plots suggested that the overall model predictions did not substantially change based on the selected ridge parameters (Supplementary Figure 2B). However, we selected $k$ = 50 as one of the smallest values at which most coefficients stabilized (neither shrinking excessively, which could lead to underfitting, nor being overly large, which could result in overfitting).

The robustness of ß estimates was tested using ten-fold cross-validation (Supplementary Figure 3A) and bootstrapping (Supplementary Figure 3B). In the ten-fold analysis, we split the training dataset into ten folds and iteratively estimated ß using nine folds for training and one for validation. This yielded 10 different ß estimates per method. For bootstrapping, we resampled the training dataset with replacement over 1000 iterations, calculating ß estimates in each run.

The in-sample scores were used to build a model to maximize coefficient stability, but the model was also tested via ten-fold cross-validation and bootstrapping. All modeling considerations and model training were based exclusively on discovery cohort. Only the final models for each level, as well as the combined model were tested on the hold-out test datasets (N = 89 and N = 21).

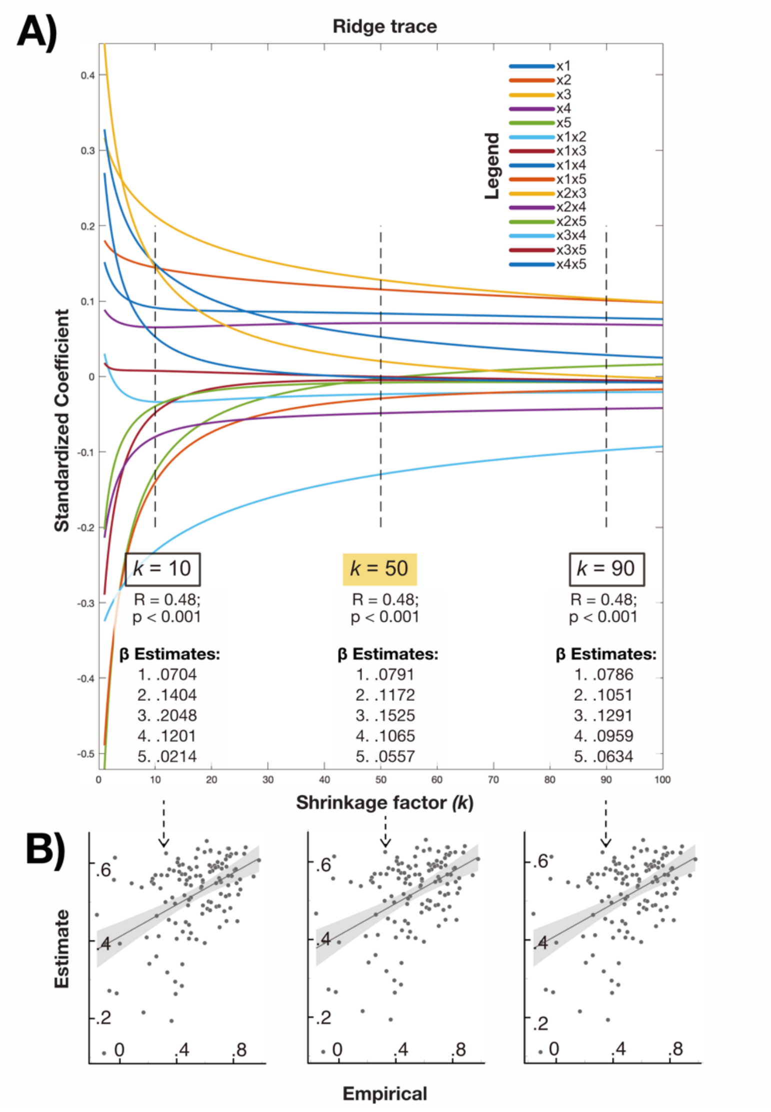

**Supplementary Figure 2. Ridge parameter (k) determination.** Five different predictors were used to fit the ridge regression. We tested 100 different values to determine the ridge parameter. A) A plot of estimates for predictors and interaction terms using 100 different ridge parameters (*k*). Three parameters were then tested to plot the ridge regression (*k* = 10, *k* = 50, *k* =90). The correlation of the predicted values and beta estimates for each of the ridge parameter tested is reported. The ridge parameter 50 (highlighted in yellow) was selected for the final model. B) Correlation plots between estimated values and the empirical values based on the ridge regression model with three different *k* parameters (*k* = 10, *k* = 50, *k* =90).

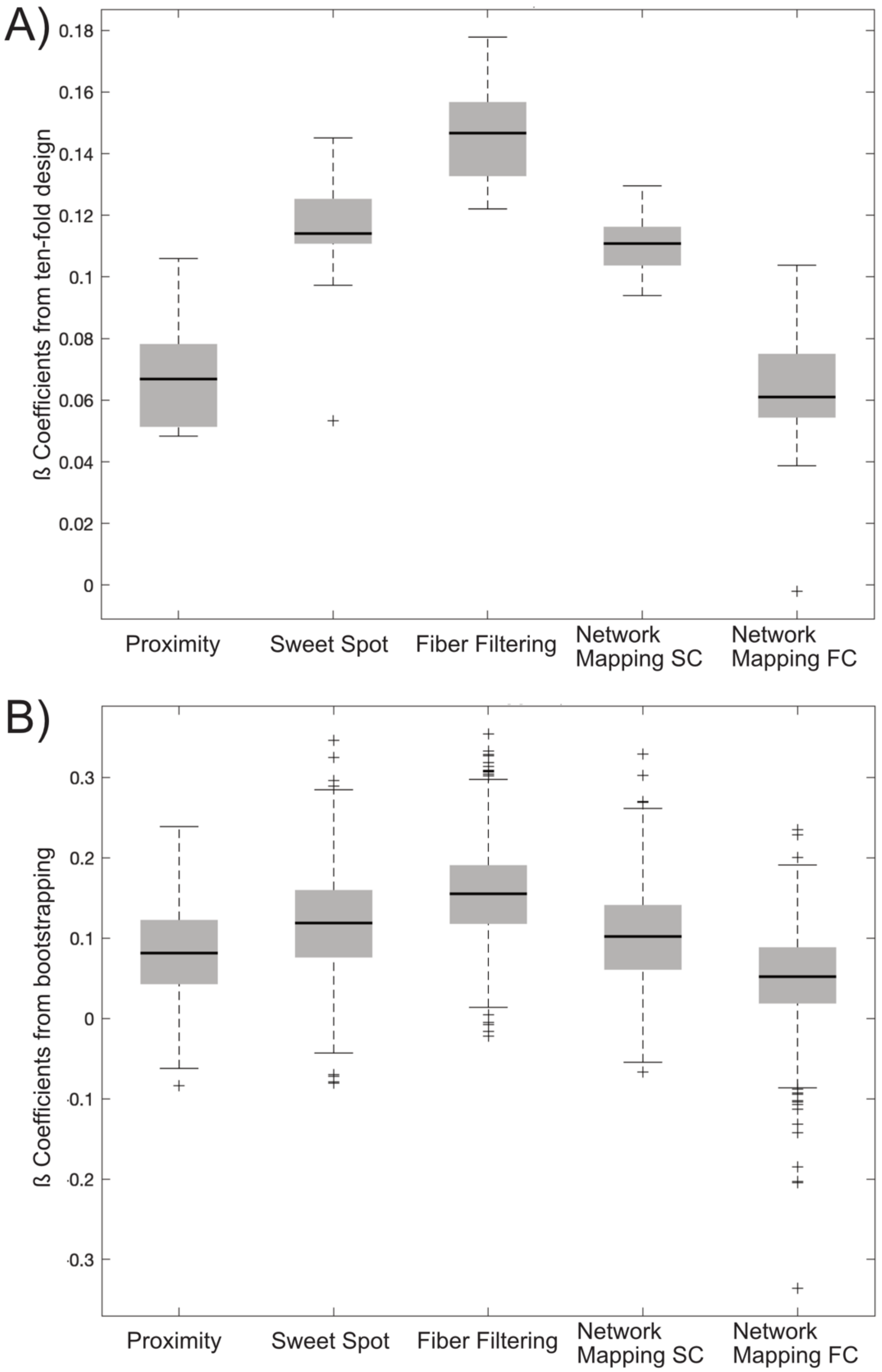

**Supplementary Figure 3. Distribution of ß estimates calculated on the training dataset using ten-fold validation and bootstrapping.** Two techniques were used to test the stability of ß estimates of our model. Black lines represent the mean, boxes represent interquartile range, whiskers represent the most extreme data points, and + sign represents the outliers. A) Our dataset was split into ten-folds and ß estimates were calculated by fitting ridge regression always on nine out of ten folds. Ten different ß estimates were calculated for each method. B) Bootstrapping with resampling method was used to calculate ß estimates in 1000 iterations. SC – structural connectivity, FC – functional connectivity.

**Supplementary Table 2**: Demographics of the discovery and test dataset

| Discovery Cohort | | | |
|---|---|---|---|
| **Center** | Berlin | Würzburg | Amsterdam |
| **Demographic information** | | | |
| **Surgical DBS Center** | Charité - Universitäts-medizin Berlin | University Hospital Würzburg | Department of Neurology, Amsterdam University Medical Center, Amsterdam, Netherlands |
| **N (female)** | 51 (17) | 44 (12) | 34 (7) |
| **Age at time of surgery (mean ± SD; range; in years)** | 60 ± 8 | 60 ± 8 | 50 ± 10 |
| **Clinical Details** | | | |
| **Average time of follow-up (in months)** | 40 | N/A | N/A |
| **UPDRS-III scores at baseline (STIM-OFF/MED-OFF) (mean ± SD)** | 38.56 ± 12.92 | 49.41 ± 12.54 | 47.03 ± 15.53 |
| **UPDRS-III scores at time of follow-up under stimulation ON condition (STIM-ON/MED-OFF) (mean ± SD)** | 20.12 ± 8.82 | 24.47 ± 10.62 | 15.07 ± 10.43 |
| **Rel. improvement (mean ± SD; in %)** | 45.34 ± 23.03 | 49.46 ± 23.97 | 64.13 ± 22.40 |
| **Abs. improvement (mean ± SD)** | 18.44 ± 11.31 | 25.37 ± 13.30 | 31.06 ± 14.97 |
| **Levodopa Response (mean % ± SD)** | 53 ± 42 | 61 ± 25 | 45 ± 34 |
| **Imaging and DBS Specification** | | | |
| **Imaging modality (post-operatively)** | MRI/CT | CT | CT |
| **Electrode models** | Medtronic 3389 | Medtronic 3389 | Medtronic 3389 |
| **Related citation** | [3,12] | [3,12] | [12] |

**Supplementary Table 3**: Information about the retrospective (Beijing & Würzburg) and prospective (Cologne) test datasets.

| Validation (Hold-Out Test) Datasets | | | |
|---|---|---|---|
| **Center** | Beijing | Würzburg | Cologne |
| | **Demographic information** | | |
| **Surgical DBS Center** | Chinese PLA General Hospital | University Hospital Würzburg | Department of Neurology, University of Cologne |
| **N (female)** | 41 (21) | 53 (19) | 24 |
| **Age at time of surgery (mean ± SD; in years)** | 61 ± 10 | 60 ± 8 | 58 ± 8 |
| | **Clinical Details** | | |
| **Time of follow-up (in months)** | N/A | 11 | N/A |
| **UPDRS-III scores at baseline (STIM-OFF/MED-OFF) (mean ± SD)** | 45.73 ± 13.12 | 42.53 ± 10.78 | 10.26 ± 3.45[a] |
| **UPDRS-III scores at time of follow-up under stimulation ON condition (STIM-ON/MED-OFF) (mean ± SD)** | 19.17 ± 10.76 | 22.45 ± 10.04 | 5 ± 3 |
| **Rel. improvement (mean ± SD; in %)** | 58.01 ± 21.29 | 46.38 ± 10.04 | 79.27 ± 14.96[b] |
| **Abs. improvement (mean ± SD)** | 26.56 ± 12.86 | 20.07 ± 10.91 | 8.07 ± 2.76[b] |
| **Levodopa Response (mean % ± SD)** | N/A | N/A | N/A |
| | **Imaging and DBS Specification** | | |
| **Imaging modality (post-operatively)** | CT | CT | CT |
| **Electrode models** | Medtronic 3389 | Boston Scientific Vercise Directed/Boston Scientific Vercise | Medtronic B3305/Boston Scientific Vercise Directed |
| **Related citation** | [12] | [12] | [22] |

[a] Only a subset of UPDRS-III items per one hemisphere was assessed, including: Upper Extremity Rigidity, Finger Tapping, Resting Tremor, Postural Tremor, and Kinetic Tremor.
[b] Absolute and relative (%) improvements in this case were calculated based on the contact with the highest clinical response.

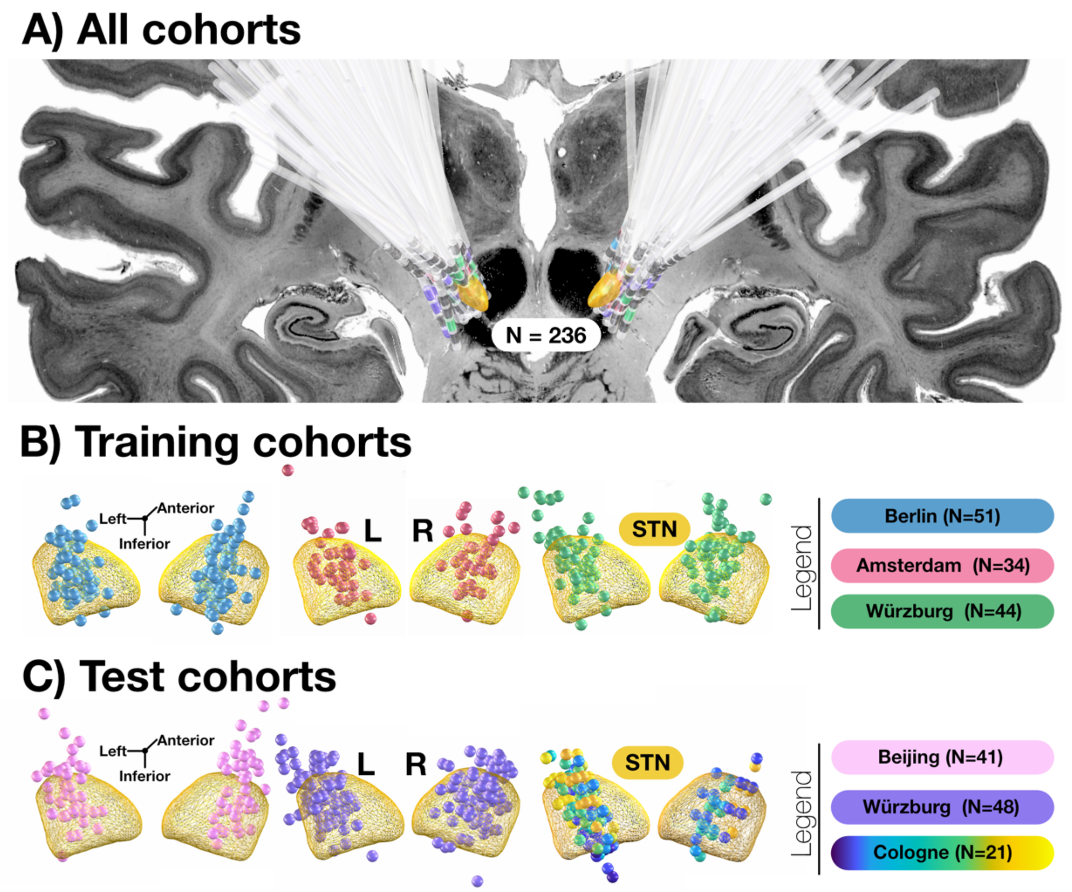

**Supplementary Figure 4. Distribution of electrode placements relative to the subthalamic nucleus (STN) across cohorts.** Electrode reconstructions across all cohorts relative to the STN (as defined by the DISTAL atlas[23]) are shown. DBS electrode placements were reconstructed using Lead-DBS software. The training cohort consisted of 129 patients who underwent bilateral DBS using omnidirectional electrodes with four stimulating contacts (Medtronic 3389), targeting the STN. The retrospective validation dataset consisted of 89 patients from two PD cohorts, implanted with three types of electrodes: Medtronic 3389 and Boston Scientific Vercise Directed. The prospectively acquired validation cohort consisted of 18 patients (21 electrodes) implanted with Boston Scientific Vercise Directed electrodes and one patient with Medtronic B3305 electrode. A) Electrodes (N = 457) from all patients included in the study are shown. A coronal slice at -20.0 mm of the BigBrain template,[24] in 100-µm resolution is visualized in the background for anatomical reference. Active contacts from patients in the training (B) and test (C) datasets are visualized as spheres relative to the STN. Since data from 8 contacts were collected for each patient in the prospectively acquired validation dataset, each patient was assigned a unique color, and the eight spheres representing their active contacts were color-coded accordingly.

### Supplementary Results 1:
### Data-driven imaging models of clinical improvement

Each of our models was compared to previously published results in the literature. At the coordinate level, our optimal target coordinate was in very close proximity to, and slightly anterior to, the one published by Caire et al., 2013 (1.37 mm distance; Supplementary Figure 5A).[25,26] At the E-field level, our sweet-spot model peaked at a similar location with the peak aligning with both the Bejjani line[27] often used for surgical targeting in clinical practice, and previously published reports (Supplementary Figure 5B).[15,28–30] At the tract level, projections from Brodmann areas (BA) 4, 6, 8 to the STN, as well as sensorimotor connections between the STN and the pallidum, and connections between the STN and the pedunculopontine nucleus (PPN), were associated with optimal clinical improvement (Supplementary Figure 5C). A table listing these connections is provided in Supplementary Table 4, where the results generally match prior reports.[12,28] At the whole-brain network level, both structural and functional DBS network mapping analyses were carried out following previously reported approaches.[3] For structural connectivity, optimal clinical improvements were associated with connections between the STN and both the SMA and pre-SMA (Supplementary Figure 5D), consistent with previously published findings.[3,15] For functional connectivity, positive connectivity to frontal and inferior parietal regions, as well as negative connectivity with the primary motor and visual cortices, were associated with optimal response (Supplementary Figure 5E), again consistent with previously published findings.[3,15]

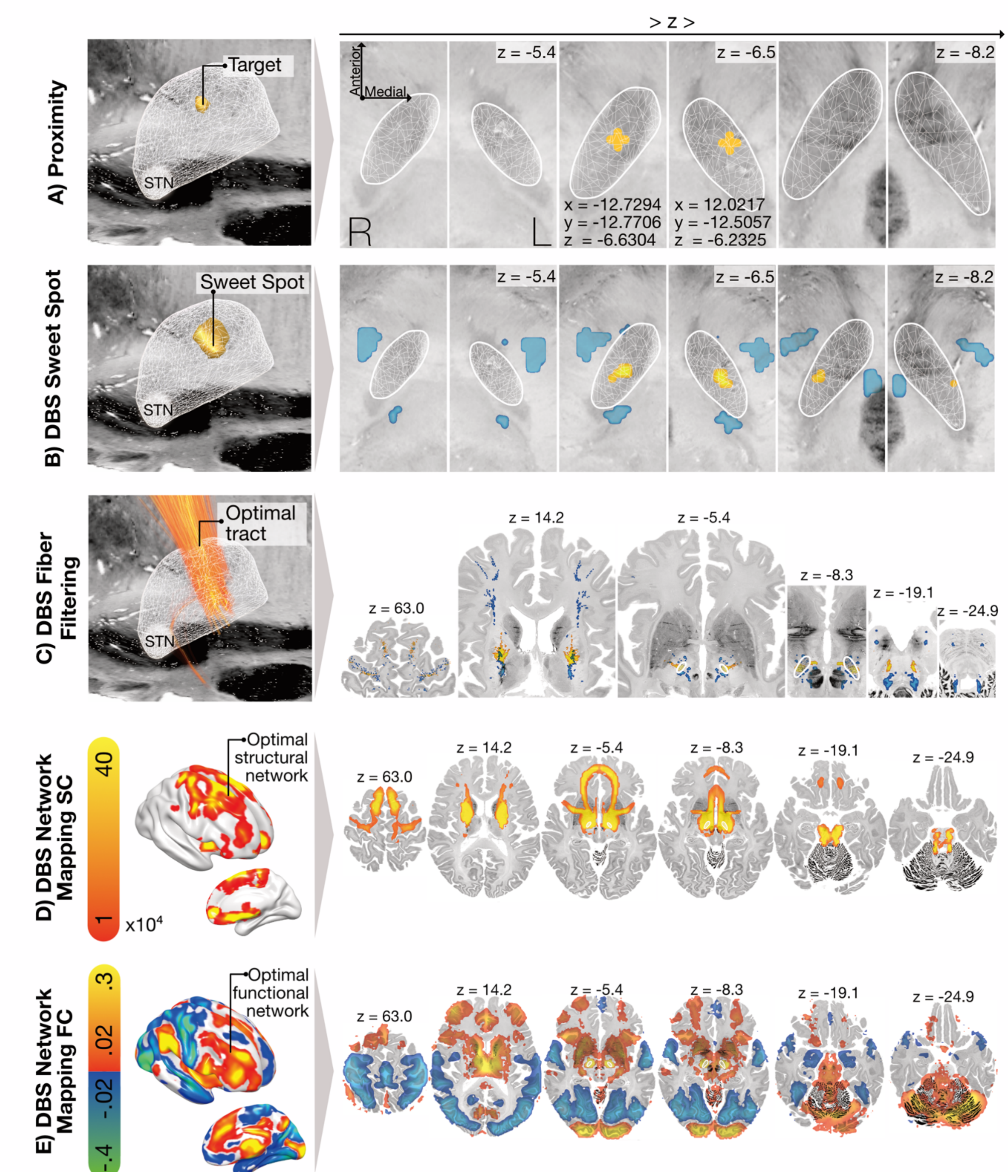

**Supplementary Figure 5. Anatomical representation of data-driven neuroimaging models of clinical improvement in Parkinson's Disease (PD) following deep brain stimulation (DBS).** Five distinct methods were used to associate clinical improvement with electrode position at different anatomical levels. For each model, the electrode position was obtained using Lead-DBS software. The BigBrain template[24] in 100-µm resolution is visualized in the background for anatomical reference. Gold indicates regions associated with clinical improvement. Blue indicates regions associated with less optimal outcomes. White mesh structures represent the subthalamic nucleus (STN). A) The optimal stimulation coordinate was defined as the weighted average of active contact locations, with weights based on clinical improvements (golden cross). Exact x, y, z coordinates are reported. B) The sweet spot was calculated using voxel-wise correlations between clinical improvements and electric field magnitudes. The resulting maps were thresholded at the uncorrected significance level ($\alpha = 0.05$). C) Optimal tract connections were calculated using a manually curated, population-based pathway atlas (Netstim Atlas V2.2) via DBS fiber filtering.[12] The optimal fiber profile was modeled by correlating peak electric field magnitudes along each fiber with clinical improvements. D) The same pathway atlas was used to calculate an optimal structural connectivity profile using a voxel-wise fashion (DBS network mapping). Voxels were assigned values based on the stimulation volumes overlapping the fibers. The combined map approach was used to determine the optimal connectivity profile, as proposed in [3]. E) The optimal functional connectivity profile was calculated from resting-state functional MRI (rs-fMRI) data from 1,087 scans in the Human Connectome Project. Seeding from the stimulation volumes, network connectivity maps were computed and correlated with clinical improvement using the same combined map approach.

**Supplementary Table 4**. DBS Fiber tracts from the Netstim Atlas V2.2 that correlated (positively and negatively) with UPDRS-III clinical improvement. The anatomical names of tracts are listed in the descending order based on their number of streamlines associated with the model.

| No. | Positive fibers | Negative fibers |
|---|---|---|
| **1.** | STN to Brodman area 6 connections | Dentatorubrothalamic tract (drtt-SMA) |
| **2.** | STN to Brodman area 4 connections | STN to Brodman area 4 connection |
| **3.** | STN to Brodman area 8 connections | STN to Brodman area 1, 2, 3 connections |
| **4.** | STN to Pallidum connection | Pedunculopontine nucleus to SMA connections |
| **5.** | STN to pedunculopontine nucleus connections | Rubrocortical tract |
| **6.** | STN to Brodman area 1, 2, 3 connections | STN to Brodman area 10 connections |
| **7.** | STN to Brodman area 10 connections | STN to Brodman area 45 and 47 connections |
| **8.** | Corticospinal tract | Corticospinal tract |
| **9.** | | Dentatorubrothalamic tract (drtt-M1) |
| **10.** | | STN to Brodman area 6 connections |
| **11.** | | STN - Pallidum |
| **12.** | | STN to Brodman area 8 connections |
| **13.** | | STN to Brodman area 24 and 32 connections |

### Supplementary Results 2:
### **Ridge regression model (additional results)**

In the joint model, DBS Fiber Filtering showed the largest standardized coefficient ($\beta = 0.152$), followed closely by DBS Sweet Spot ($\beta = 0.117$), structural DBS Network Mapping ($\beta = 0.106$), coordinate proximity ($\beta = 0.079$) and functional DBS Network Mapping ($\beta = 0.056$) (Figure 3B). Statistical significance of the estimates is typically not reported in ridge regression models due to the shrinkage factor, that influences the ß estimates determined by the model.

Our joint model explained 11% of the variance in clinical outcomes in the heterogeneous, group-level validation cohort. As indicated by the coefficient of determination, the variance explained by fitting a general linear model to surrogate variables from individual methods on the validation cohort was lower in all instances ($R^2$ proximity = 0.037; $R^2$ DBS Sweet Spot = 0.066; $R^2$ DBS Fiber Filtering = -0.030; $R^2$ DBS Network Mapping SC = 0.092; $R^2$ DBS Network Mapping FC = 0.089).

Due to different clinical improvement scale of the prospectively acquired contact-wise stimulation cohort, calculating actual clinical predictions was not possible. However, we fitted general linear model for each method and correlated the estimates with the empirical clinical improvement (Supplementary Figure 6). For each method, we calculated the success rate (%) of matching the top empirical contact with the one selected by our model, as well as the empirical contact ranking being top 2 or top 3 contacts selected by our model. We also calculated the success rate (%) of our model in selecting the correct segment for beneficial stimulation (Supplementary Table 5).

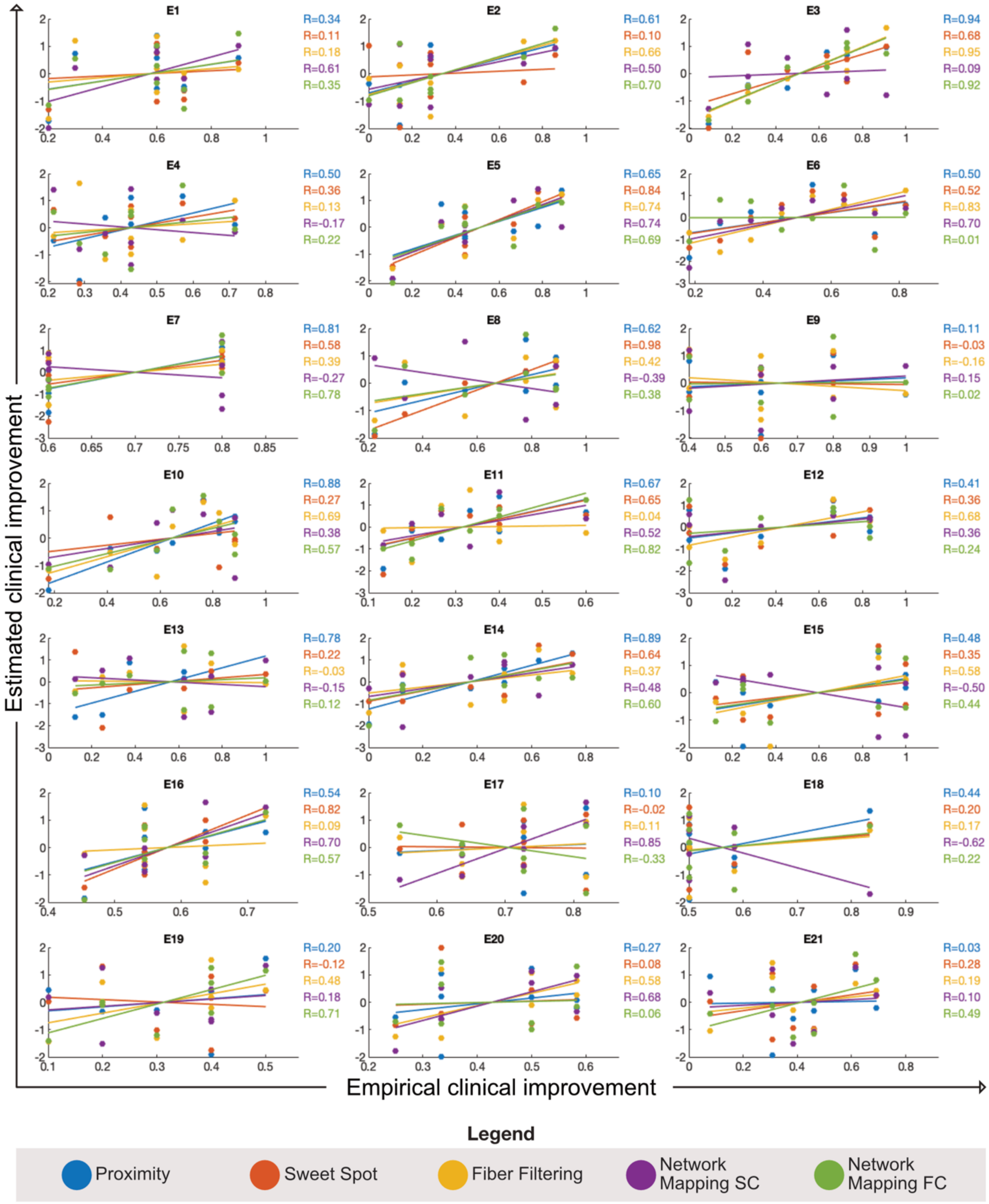

**Supplementary Figure 6. Estimates of clinical improvement in the contact-wise stimulation cohort using five different modelling methods.** Each model was trained on one independent dataset. A general linear model was then fitted to that dataset, and clinical improvements were estimated in the new contact-wise stimulation cohort. Estimated improvements for each method were normalized and plotted on the y-axis against the empirical clinical improvement associated with each electrode contact on the x-axis. Note that in some cases, multiple electrode contacts shared the same empirical improvement value.

**Supplementary Table 5. Individual model performance on contact-wise stimulation data.** The percentage (%) of matches between the optimal clinical contact and the model-selected top contact, top two contacts, top three contacts, as well as the agreement between the model-selected and clinically selected stimulation levels across different methods.

| | **Top 1 Contacts (%)** | **Top 2 Contacts (%)** | **Top 3 Contacts (%)** | **Level (%)** |
|---|---|---|---|---|
| **Proximity** | 38 | 62 | 71 | 71 |
| **Sweet Spot** | 24 | 48 | 67 | 52 |
| **Fiber Filtering** | 24 | 48 | 76 | 57 |
| **Network Mapping SC** | 19 | 43 | 62 | 52 |
| **Network Mapping FC** | 29 | 52 | 67 | 57 |
| **Overall Model** | 38 | 57 | 81 | 71 |

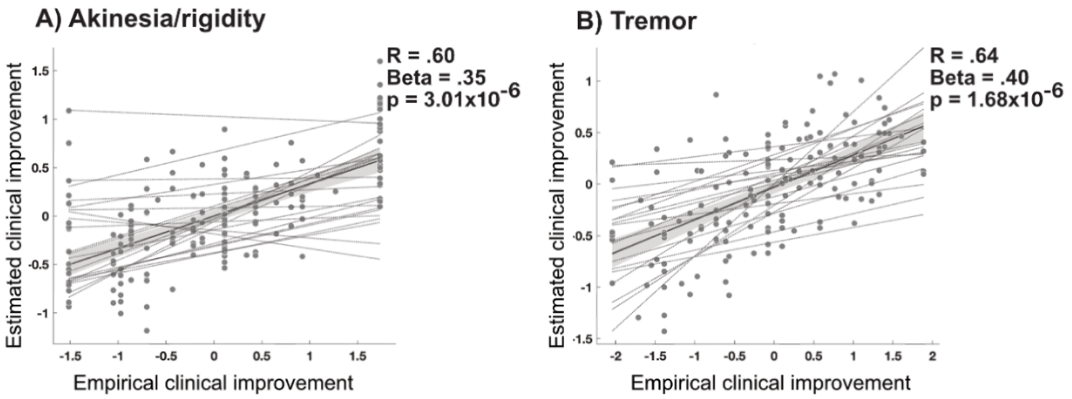

**Supplementary Figure 7. Linear mixed-effect model results for akinetic and tremor symptoms.** The combined ridge regression model was used to estimate clinical improvements for individual contacts specific to electrode placements in 21 cases. A linear mixed-effect model was fitted, with random intercepts for each individual electrode. A) Model-derived estimates of clinical improvement were positively correlated with the empirical improvements in akinesia. B) Model-derived estimates of clinical improvement were also positively correlated with the empirical improvement in tremor.

### Supplementary References

1. Cavallieri F, Fraix V, Bove F, et al. Predictors of Long-Term Outcome of Subthalamic Stimulation in Parkinson Disease. *Annals of Neurology*. 2020;89(3):587-597. doi:10.1002/ana.25994

2. Soulas T, Sultan S, Gurruchaga JM, Palfi S, Fénelon G. Depression and Coping as Predictors of Change After Deep Brain Stimulation in Parkinson's Disease. *World Neurosurgery*. 2011;75(3):525-532. doi:10.1016/j.wneu.2010.06.015

3. Horn A, Reich M, Vorwerk J, et al. Connectivity Predicts deep brain stimulation outcome in Parkinson disease: DBS Outcome in PD. *Ann Neurol*. 2017;82(1):67-78. doi:10.1002/ana.24974

4. Alonso F, Zsigmond P, Wårdell K. Influence of Virchow-Robin spaces on the electric field distribution in subthalamic nucleus deep brain stimulation. *Clinical Neurology and Neurosurgery*. 2021;204:106596. doi:10.1016/j.clineuro.2021.106596

5. Muthuraman M, Deuschl G, Koirala N, Riedel C, Volkmann J, Groppa S. Effects of DBS in parkinsonian patients depend on the structural integrity of frontal cortex. *Sci Rep*. 2017;7(1):43571. doi:10.1038/srep43571

6. Richards M, Marder K, Cote L, Mayeux R. Interrater reliability of the unified Parkinson's disease rating scale motor examination. *Movement Disorders*. 1994;9(1):89-91. doi:10.1002/mds.870090114

7. Bennett DA, Shannon KM, Beckett LA, Goetz CG, Wilson RS. Metric properties of nurses' ratings of parkinsonian signs with a modified Unified Parkinson's Disease Rating Scale. *Neurology*. 1997;49(6):1580-1587. doi:10.1212/WNL.49.6.1580

8. Siderowf A, McDermott M, Kieburtz K, et al. Test-Retest reliability of the Unified Parkinson's Disease Rating Scale in patients with early Parkinson's disease: Results from a multicenter clinical trial. *Mov Disord*. 2002;17(4):758-763. doi:10.1002/mds.10011

9. Metman LV, Myre B, Verwey N, et al. Test-retest reliability of UPDRS-III, dyskinesia scales, and timed motor tests in patients with advanced Parkinson's disease: An argument against multiple baseline assessments. *Mov Disord*. 2004;19(9):1079-1084. doi:10.1002/mds.20101

10. Maks CB, Butson CR, Walter BL, Vitek JL, McIntyre CC. Deep brain stimulation activation volumes and their association with neurophysiological mapping and therapeutic outcomes. *J Neurol Neurosurg Psychiatry*. 2009;80(6):659-666. doi:10.1136/jnnp.2007.126219

11. Noecker AM, Frankemolle-Gilbert AM, Howell B, et al. StimVision v2: Examples and Applications in Subthalamic Deep Brain Stimulation for Parkinson's Disease. *Neuromodulation*. 2021;24(2):248-258. doi:10.1111/ner.13350

12. Rajamani N, Friedrich H, Butenko K, et al. Deep brain stimulation of symptom-specific networks in Parkinson's disease. *Nat Commun*. 2024;15(1):4662. doi:10.1038/s41467-024-48731-1

13. Neudorfer C, Butenko K, Oxenford S, et al. Lead-DBS v3.0: Mapping deep brain stimulation effects to local anatomy and global networks. *NeuroImage*. 2023;268:119862. doi:10.1016/j.neuroimage.2023.119862

14. Meyer GM, Hollunder B, Li N, et al. Deep Brain Stimulation for Obsessive-Compulsive Disorder: Optimal Stimulation Sites. *Biological Psychiatry*. 2024;96(2):101-113. doi:10.1016/j.biopsych.2023.12.010

15. Horn A, Li N, Dembek TA, et al. Lead-DBS v2: Towards a comprehensive pipeline for deep brain stimulation imaging. *NeuroImage*. 2019;184:293-316. doi:10.1016/j.neuroimage.2018.08.068

16. Sobesky L, Goede L, Odekerken VJJ, et al. Subthalamic and pallidal deep brain stimulation: are we modulating the same network? *Brain*. 2022;145(1):251-262. doi:10.1093/brain/awab258

17. Åström M, Diczfalusy E, Martens H, Wårdell K. Relationship between neural activation and electric field distribution during deep brain stimulation. *IEEE Trans Biomed Eng*. 2015;62(2):664-672. doi:10.1109/TBME.2014.2363494

18. Van Essen DC, Ugurbil K, Auerbach E, et al. The Human Connectome Project: A data acquisition perspective. *Neuroimage*. 2012;62(4):2222-2231. doi:10.1016/j.neuroimage.2012.02.018

19. Goede LL, Al-Fatly B, Li N, et al. Convergent mapping of a tremor treatment network. *Nat Commun*. 2025;16(1):4772. doi:10.1038/s41467-025-60089-6

20. Ríos AS, Oxenford S, Neudorfer C, et al. Optimal deep brain stimulation sites and networks for stimulation of the fornix in Alzheimer's disease. *Nat Commun*. 2022;13(1):7707. doi:10.1038/s41467-022-34510-3

21. Horn A, Reich MM, Ewert S, et al. Optimal deep brain stimulation sites and networks for cervical vs. generalized dystonia. *Proc Natl Acad Sci USA*. 2022;119(14):e2114985119. doi:10.1073/pnas.2114985119

22. van der Linden C, Berger T, Brandt GA, et al. Accelerometric Classification of Resting and Postural Tremor Amplitude. *Sensors (Basel)*. 2023;23(20):8621. doi:10.3390/s23208621

23. Ewert S, Plettig P, Li N, et al. Toward defining deep brain stimulation targets in MNI space: A subcortical atlas based on multimodal MRI, histology and structural connectivity. *NeuroImage*. 2018;170:271-282. doi:10.1016/j.neuroimage.2017.05.015

24. Amunts K, Lepage C, Borgeat L, et al. BigBrain: An Ultrahigh-Resolution 3D Human Brain Model. *Science*. 2013;340(6139):1472-1475. doi:10.1126/science.1235381

25. Caire F, Ranoux D, Guehl D, Burbaud P, Cuny E. A systematic review of studies on anatomical position of electrode contacts used for chronic subthalamic stimulation in Parkinson's disease. *Acta Neurochir (Wien)*. 2013;155(9):1647-54-discussion 1654. doi:10.1007/s00701-013-1782-1

26. Horn A, Kühn AA, Merkl A, Shih L, Alterman R, Fox M. Probabilistic conversion of neurosurgical DBS electrode coordinates into MNI space. *NeuroImage*. 2017;150:395-404. doi:10.1016/j.neuroimage.2017.02.004

27. Bejjani BP, Dormont D, Pidoux B, et al. Bilateral subthalamic stimulation for Parkinson's disease by using three-dimensional stereotactic magnetic resonance imaging and electrophysiological guidance. *Journal of Neurosurgery*. 2000;92(4):615-625. doi:10.3171/jns.2000.92.4.0615

28. Akram H, Sotiropoulos SN, Jbabdi S, et al. Subthalamic deep brain stimulation sweet spots and hyperdirect cortical connectivity in Parkinson's disease. *NeuroImage*. 2017;158:332-345. doi:10.1016/j.neuroimage.2017.07.012

29. Bot M, Schuurman PR, Odekerken VJJ, et al. Deep brain stimulation for Parkinson's disease: defining the optimal location within the subthalamic nucleus. *J Neurol Neurosurg Psychiatr*. Published online January 20, 2018:jnnp-2017-316907-7. doi:10.1136/jnnp-2017-316907

30. Horn A. The impact of modern-day neuroimaging on the field of deep brain stimulation. *Current Opinion in Neurology*. 2019;32(4):511-520. doi:10.1097/WCO.0000000000000679
